# Heterogeneous SARS-CoV-2 kinetics due to variable timing and intensity of immune responses

**DOI:** 10.1101/2023.08.20.23294350

**Authors:** Katherine Owens, Shadisadat Esmaeili-Wellman, Joshua T. Schiffer

**Affiliations:** Fred Hutchinson Cancer Center, Vaccine and Infectious Diseases Division; University of Washington, Department of Medicine

## Abstract

The viral kinetics of documented SARS-CoV-2 infections exhibit a high degree of inter-individual variability. We identified six distinct viral shedding patterns, which differed according to peak viral load, duration, expansion rate and clearance rate, by clustering data from 768 infections in the National Basketball Association cohort. Omicron variant infections in previously vaccinated individuals generally led to lower cumulative shedding levels of SARS-CoV-2 than other scenarios. We then developed a mechanistic mathematical model that recapitulated 1510 observed viral trajectories, including viral rebound and cases of reinfection. Lower peak viral loads were explained by a more rapid and sustained transition of susceptible cells to a refractory state during infection, as well as an earlier and more potent late, cytolytic immune response. Our results suggest that viral elimination occurs more rapidly during omicron infection, following vaccination, and following re-infection due to enhanced innate and acquired immune responses. Because viral load has been linked with COVID-19 severity and transmission risk, our model provides a framework for understanding the wide range of observed SARS-CoV-2 infection outcomes.

## Introduction

COVID-19 public health emergency status has lapsed in the United States, but community levels of severe acute respiratory syndrome coronavirus 2 (SARS-CoV-2) remain significant (https://covid.cdc.gov/covid-data-tracker/#datatracker-home). SARS-CoV-2 immunity in the population is now highly heterogeneous due to varying degrees of prior infection and vaccination^1^. Also, successive circulating SARS-CoV-2 variants of concern (VOC) with different immune evasion and infectivity properties continue to emerge. This has resulted in a wider variability of viral shedding patterns than those observed during infection with the ancestral strain in the early months of 2020^2, 3^. Understanding the heterogeneous upper respiratory tract (URT) kinetics of SARS-CoV-2 enables informed design of health interventions such as testing, isolation, quarantine, and drug therapies.

Mathematical models are a vital tool for understanding mechanisms underlying observed patterns of viral expansion and clearance^4–9^. To date, studies fitting SARS-CoV-2 dynamic models to viral load trajectories have estimated the timing of innate and acquired immune responses and predicted transmission parameters, including super-spreader events^10–22^. These models facilitated estimates of key quantities such as expected duration of the infectious period and the timing of peak viral load relative to symptom onset^20, 23–25^. They also provided a theoretical means for testing treatment regimens and predicted that treatment within 5 days of symptom onset would likely be associated with higher efficacy^11, 22, 24, 26, 27^, an outcome that has since been verified in multiple clinical trials^28–30^. These models were also the first to suggest that viral rebound may occur in the context of early antiviral treatments^11^.

However, early modeling studies only considered data from a small number of infected individuals^11, 20, 22–27, 31–35^, and often drew either entirely from previously uninfected and/or unvaccinated cohorts^13^. Another consistent limitation was that most available data did not capture early timepoints during the pre-symptomatic phase of infection. Model results are therefore, not easily generalized to current SARS-CoV-2 conditions.

The National Basketball Association’s (NBA) daily testing program occurred regardless of symptoms and identified 2,875 infections between June 2020 and January 2022, spanning the alpha, delta, and early omicron VOC waves, as well as the roll-out of vaccines and boosters. Hay et al. used a statistical approach to quantify the impact of immune history and variant on SARS-CoV-2 viral kinetics and infection rebound in this data set^36^. However, a more mechanistic modeling approach is required to understand observed kinetic variability in this cohort.

Here, we identify six distinct shedding patterns in the NBA cohort data. We then compare how well candidate models which extend the classical target-cell limited model previously published by Goyal et al.^11, 22^ and Ke at al.^20, 23^ recapitulate the longitudinal upper respiratory tract (URT) viral load data from 1510 sufficiently documented infections. After obtaining data-validated parameter estimates for each individual infection, we identify the factors underlying differing rates of viral expansion and clearance, peak viral loads, and duration of infection observed in the data. We use the model to identify differences between the timing and intensity of the immune response during initial and re-infections and identify a potential explanation for viral rebound observed in the cohort.

## Results

### Viral shedding kinetics according to SARS-CoV-2 VOC

We first analyzed viral kinetics observed in the cohort according to VOC. For pre-VOC, alpha, delta, and omicron variants, we observed variable kinetics among cohort participants. Median values differed between variants, with omicron variant having slightly lower peak viral loads and earlier clearance, while delta had the highest peak viral loads and pre-VOC had the longest time to clearance (**Fig 1a-d**). A high proportion of the infections caused by omicron variants occurred in participants who had received either two or three vaccine doses, whereas pre-delta infections mostly occurred in unvaccinated individuals (**Fig 1e**).

**Figure 1:**
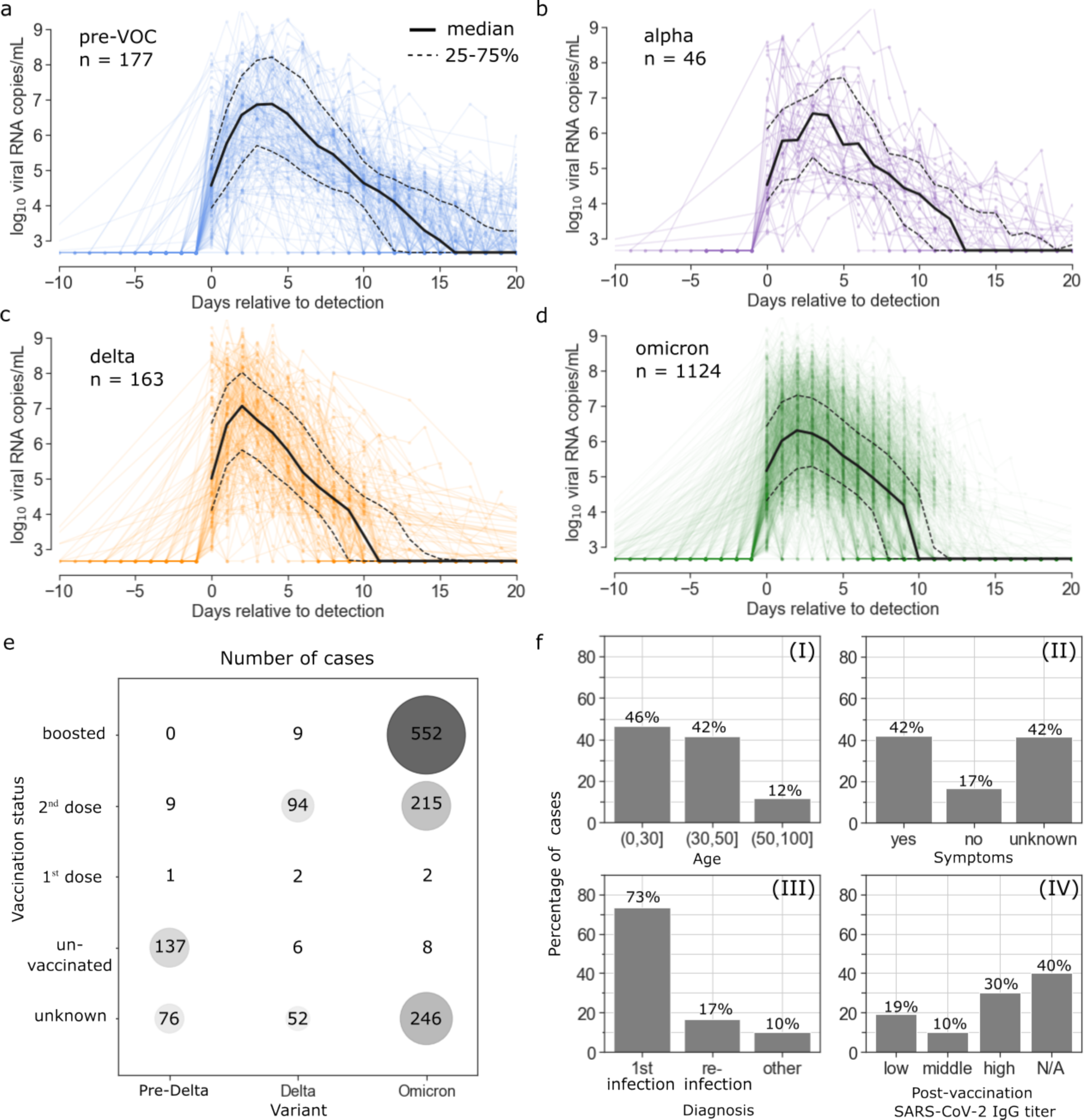
Viral kinetics by variant in the National Basketball Association cohort from June 2020-January 2022. 1510 SARS-CoV-2 infections are documented. Time series are stratified by variant with individual viral loads plotted in color, the median viral load plotted with a solid black line, and the 25^th^ and 75^th^ percentiles plotted in dashed black lines for (a) pre-variant of concern viruses, (b) alpha, (c) delta, and (d) omicron infections. (e) Bubble plot showing the relationship between variant of infection and vaccination status of the individual. Both the color and the size of the circle indicate the number of infections in each category. (f) Additional information about infections includes age, presence of symptoms, re-infection status, and pre-infection antibody titer following vaccination.

The age structure of the NBA cohort differs significantly from the general population. Of the cases documented, 46% occurred in individuals under the age of 30, 42% occurred in individuals between the ages of 30 and 50, and only 12% occurred in individuals over the age of 50 (**Fig 1f**). Symptom status was noted for 59% of infections, of which 71% were symptomatic (**Fig 1f**). The level of post-vaccination, pre-infection SARS-CoV-2 IgG was measured in 60% of infections. When stratifying patients into tertiles, Hay et al. identified low antibody titers as less than 125 (arbitrary units [AU]/ml), mid-range titers as greater than 125 AU but less than 250 AU, and high titers as greater than 250 AU with the most infections occurring in the highest tertile (**Fig 1f**). 17% of observed infections were reinfections of individuals followed longitudinally (**Fig 1f**).

### Six distinct SARS-CoV-2 shedding patterns

We identified a subset of infections in the NBA cohort as “well-documented” if they had at least 4 quantitative positive viral load measurements starting within 5 days of detection, and if infection was documented for 3 weeks, or viral elimination was confirmed with 2 sequential negative test results. This reduced the data set to 810 well-documented infections. To eliminate intra-individual variability from this data set, we retained one infection from individuals with multiple documented infections further narrowing our focus to 768 infections. We then applied k-means clustering to the viral load data, clustering infections into 6 distinct viral shedding patterns (**Fig. 2a-c**) which differed according to time to viral elimination (**Fig. 2c,d**), area under the viral curve (**Fig. 2c,e**), peak viral load (**Fig. 2c,f**) and time to peak (**Fig. 2c,g**).

**Figure 2:**
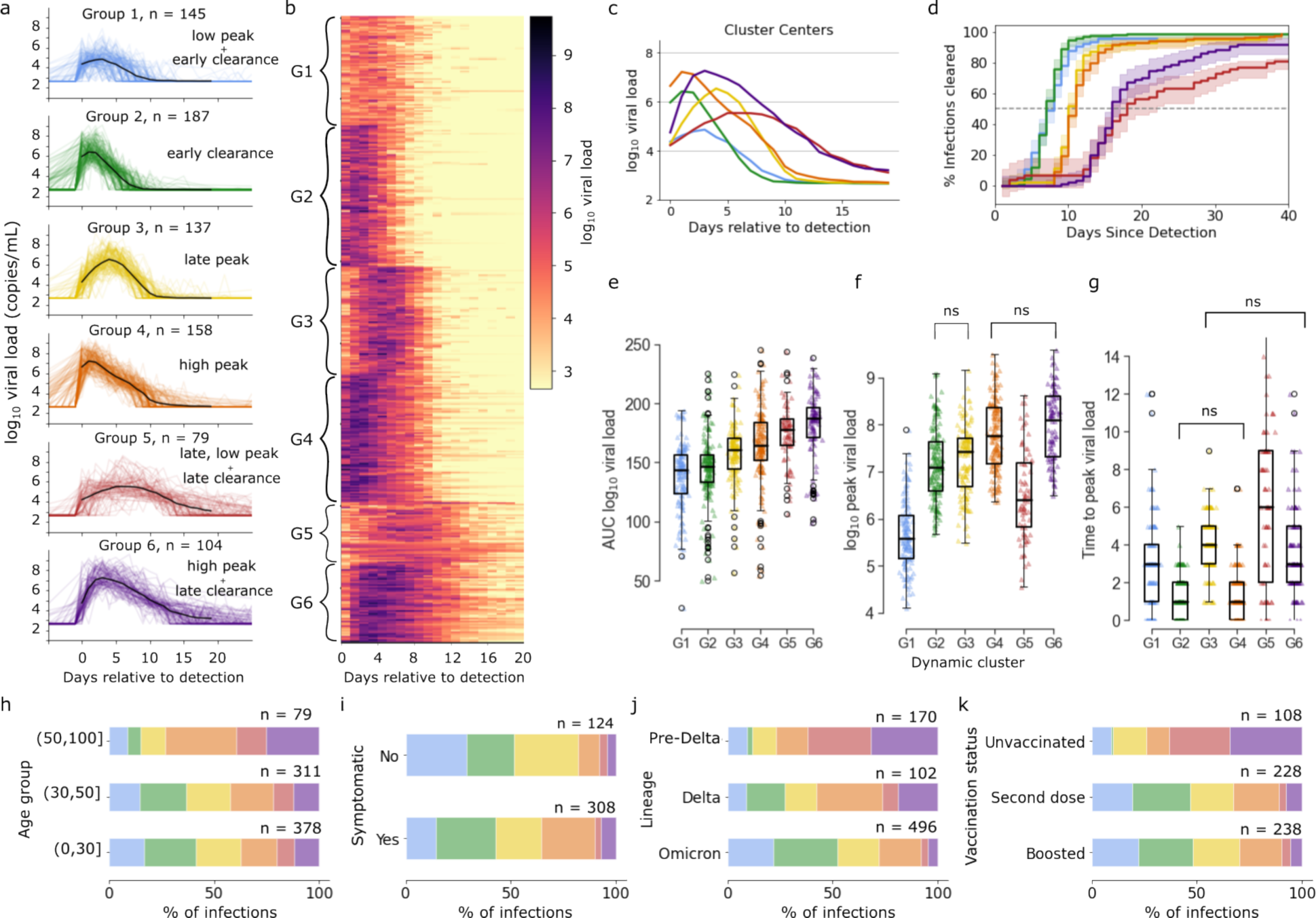
Distinct viral dynamic profiles in the National Basketball Association cohort from June 2020-January 2022. (a) Trajectories stratified by cluster assignment after k-means clustering with k = 6. Cluster centers are shown in black. (b) Heat map of log viral load over time. Each row corresponds to an infection and trajectories are ordered according to cluster. (c) Cluster centers plotted on the same axis demonstrate differing peak viral loads, time of viral peak, clearance rate and time to clearance by cluster. (d) The proportion of infections cleared over time for each cluster with 95% confidence interval shaded. Boxplots of (e) area under the log_10_ viral load curve, (f) peak viral load for different dynamic groups, and (g) days between detection and peak viral load. According to a Mann-Whitney U-test, distinctions in the mean for all possible pairs of groups are significant (p_adjusted_ < .05) except for the pairs marked “ns.” In the final row, stacked bar charts indicate the percentage of cases that fall into each dynamic group when cases are stratified by (h) age group, (i) symptom status, (j) infecting variant, and (k) vaccination status.

The first group had low peak viral loads and early median time to clearance (**Fig. 2a-g**). The second group had a slightly earlier and significantly higher peak than group 1, but similarly short duration (**Fig 2a-g**). The third group had a similar peak viral load compared to group 2, but with a longer time to peak viral load and later clearance (**Fig 2a-g**). The fourth group had the fastest expansion rate, reaching a high, early peak viral load, but maintaining similar median time to clearance as group 3 (**Fig 2a-g**). The fifth group had the slowest expansion rate, taking the longest time to reach the second lowest peak viral load and had the longest median time to clearance among the groups (**Fig 2a-g**). In contrast with the prolonged low-level shedding of group 5, the sixth group had high, somewhat early peak and a long shedding duration (**Fig 2a-g**).

The proportion of cases that fell into each dynamic group varied when we stratified by characteristics included in the data set. The dynamic groups with highest AUC, groups 5 and 6, made up 39% of the infections in the 50 plus age group, whereas 21% of infections in the under 30 group were in the high AUC groups (**Fig. 2h**). Among confirmed asymptomatic infections, 29% of cases fell into group 1, defined by low peak and early time to clearance, relative to only 14% of confirmed symptomatic cases; the slowly expanding group 3 cases were also less likely to be symptomatic while high, early peak group 4 cases were more often symptomatic (**Fig. 2i**). High AUC shedding patterns were also more prevalent among infections with SARS-CoV-2 variants from earlier in the pandemic, making up 62% of pre-VOC infections, 27% of delta infections, and only 8% of omicron infections (**Fig. 2j**). Amongst unvaccinated individuals, high AUC infection patterns were much more frequent—63% of infections in unvaccinated individuals fell into groups 5 and 6, compared with 11% and 9% of infections for those whose most recent SARS-CoV-2 vaccine was their second dose or booster respectively (**Fig. 2k**).

### Mathematical model fit to viral loads from 1510 SARS-CoV-2 infections

To identify factors underlying the varied viral shedding patterns in the NBA cohort, we developed competing mechanistic mathematical models of viral and immune dynamics and selected the best model according to data-fitting criteria. The most complex model tested adapts previously published ordinary differential equations models for within-host SARS-CoV-2 infections by combining elements introduced by Goyal et al.^11, 22^ and Ke et al.^20, 23^. For this model we made mechanistic assumptions inherent to many pre-existing viral dynamic models including a viral load dependent infectivity, viral production by infected cells, a limited number of susceptible cells, and a pre-production eclipse phase for infected cells. The possible immune mechanisms included in the model were conversion of susceptible cells to an infection-refractory state dependent on the number of infected cells (presumably representing innate responses to infection), density-dependent death of infected cells as a proxy for an intensifying cytolytic innate response to a higher burden of infection, and a delayed cytolytic acquired immune response (**Fig. 3a; Materials and Methods**).

**Figure 3:**
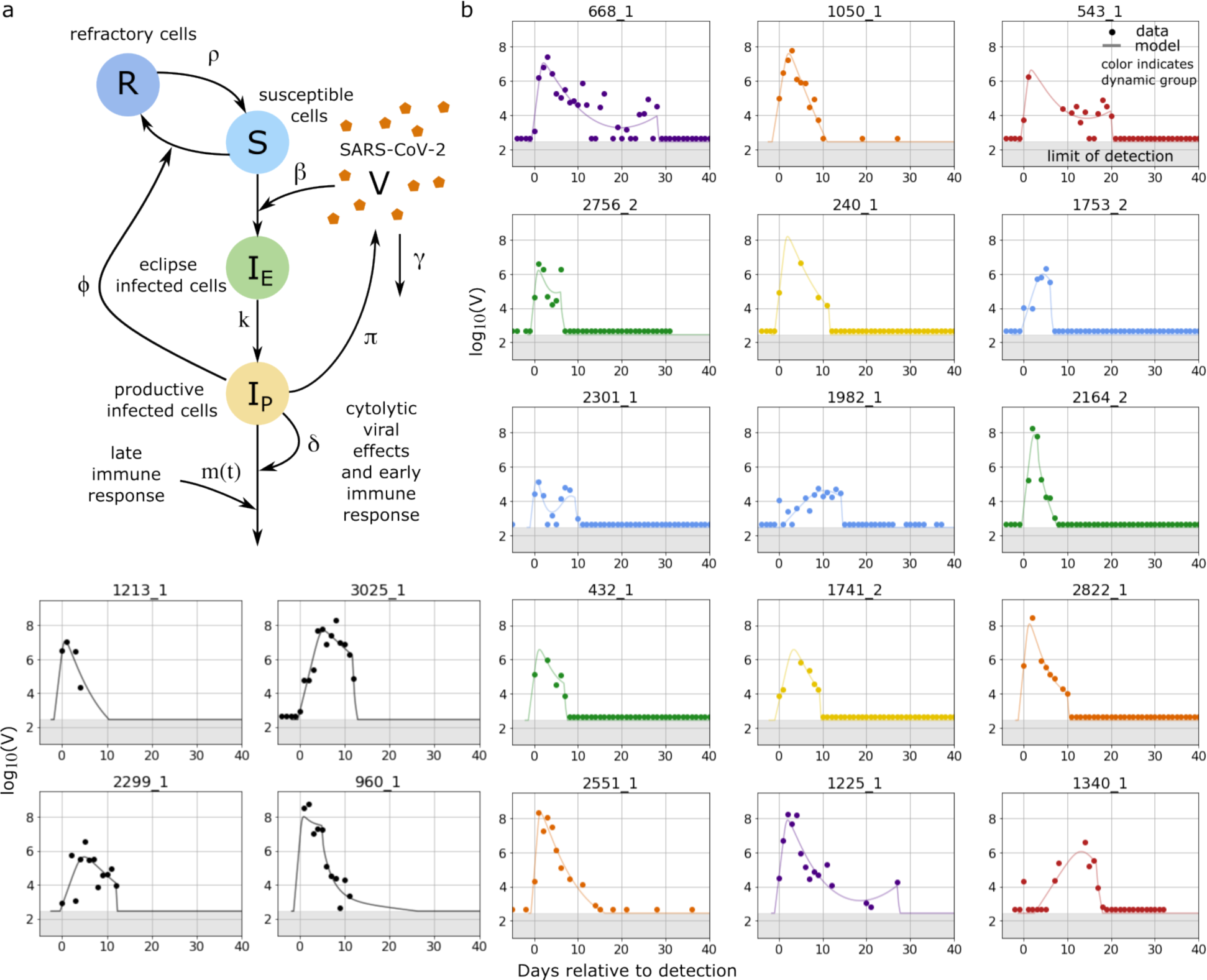
Mechanistic mathematical model with fits to viral loads from each cluster. (a) Schematic of the ordinary differential equations model used to simulate SARS-CoV-2 infection with state variables indicated by capital letters, interactions indicated by arrows and parameters indicated by symbols adjacent to arrows. The model contains an early and late cytolytic immune response. (b) Examples of data from individual infections and corresponding model simulations colored according to cluster identified via k-means clustering as in **Fig 2** with group 1 in blue, group 2 in green, group 3 in yellow, group 4 in orange, and group 5 in red and group 6 in purple. The black examples were not included in cluster analysis. The model also captures instances of rebound or non-monotonic clearance.

We used a nonlinear, mixed-effects framework to estimate model parameters for the 1510 infections documented in 1442 individuals in the NBA cohort that had at least 4 quantitative viral load measurements (**Materials and Methods**). We first used a representative subsample of these infections to compare model fits for the full model, illustrated in **Fig. 3a** and written out in equation (1), and reasonable simplifications, in which one or more immune mechanism was removed (**Materials and Methods, Table S1**). Under model selection criteria that balance simplicity with accuracy, the best model to explain the NBA data was the full model except for the density-dependent death of infected cells. This model has been previously studied by Ke et al.^20^ We then refit the best model to all infections. It is possible that models outside of the collection tested here could describe the data better; however, the fits that we achieve with this model were highly accurate for most members of the cohort from all 6 shedding clusters (**Fig. 3b, Fig. S6**).

### Differences in timing and intensity of immune response as an explanation for heterogeneous shedding patterns

We next sought to explore possible virologic and immunologic explanations for different observed viral shedding patterns. For relevant model quantities, we calculated the mean within each dynamic group at each time point and a 95% confidence interval, assuming normally distributed values. Mean viral loads projected by the model for each group (**Fig. 4a**) resembled those from the actual data (**Fig 2c**). Quantitative kinetic features extracted from model simulation output including peak viral load **(Fig S1a)**, time to peak **(Fig S1b)**, viral area under the curve **(Fig S1c),** and shedding kinetic group also agreed well with those extracted from the cohort data (**Fig S1d-g**). Projections for susceptible cells and infected cells suggest dynamics which track closely to viral load that differ accordingly among shedding subgroups **(Fig. 4b,c).**

**Figure 4:**
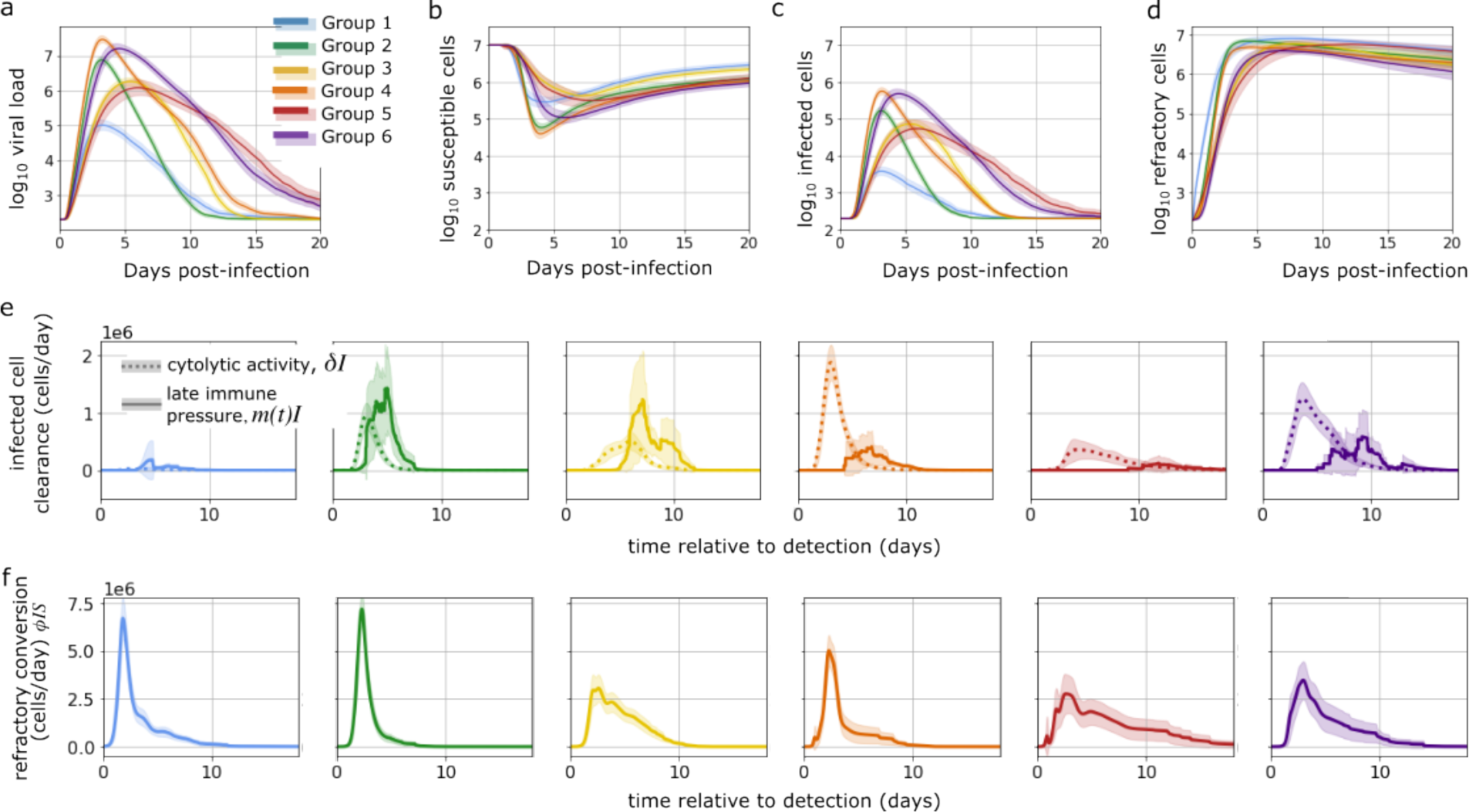
Mechanistic differences between dynamic groups. Panels (a-d) show the mean viral load and cell populations in the mechanistic model for each group over time with 95% confidence interval shaded. The quantities are (a) log viral load, log_10_(V), (b) number of susceptible cells, log_10_(S), (c) number of active infected cells, log_10_(I) and (d) the number of cells refractory to infection log_10_(R). Next, we plot the mean and standard deviation of immune pressures over time for each dynamic group. Panel (e) shows the infected cell clearance due to both constant cytolytic activity and delayed immune pressure. Panel (f) shows the conversion of susceptible cells to a refractory state.

To delineate mechanistic drivers of shedding variability, we calculated the Pearson correlation coefficient between individual estimates for each model parameter and 4 viral kinetic quantities predicted by the mathematical model: log of peak viral load, time to peak viral load, shedding duration, and area under the log viral load curve (**Fig. S2a-d**). Peak viral load correlated strongly with viral production rate, *π*, and had a strong inverse correlation with the rate of conversion of susceptible cells to a refractory state, *ϕ* (**Fig. S2a**). A linear model mapping log(*π*/*ϕ*) to log peak viral load predicted by the model explained a large amount of variability (**Fig. S2e**). The timing of peak viral load inversely correlated strongly with *π*, *β*, and *ϕ* (**Fig. S2b**). We fit an exponential model for time to peak viral load relative to infection as a function of log_10_(*βπ*), which again explained a large amount of observed variability, *R*^2^ = 0.76 (**Fig. S2f**).

Shedding duration correlated most strongly with the time of onset of acquired immunity in the model, *τ* (**Fig. S2c**). Overall, the value of *τ* did not predict the time of clearance very well. This is because for a significant number of individuals particularly in groups 1-4, acquired immunity was established after the virus was already cleared **(Fig S2g)**. In groups 5 and 6, timing of acquired immunity onset was more predictive of shedding duration (*R*^2^ = 0.61) because acquired immunity was usually established before the virus was cleared. Numerous model parameters influenced viral AUC though *τ* and *ϕ* were most important **(Fig S2d).**

The viral shedding pattern for group 1 was notable for a low peak and early clearance of infection **(Fig S2h)**. These mild virologic outcomes occurred due to rapid generation of refractory cells. Early onset of acquired immune pressure was only occasionally necessary for viral clearance (**Fig. 4d-f, Fig S2j**). The higher viral peak in group 2 was driven by relatively higher values of viral production and viral infectivity and low conversion to a refractory state (**Fig. S2j-l**). In group 2 infections, innate and acquired immunity both play a role in the clearance of infected cells (**Fig. 4e**). Infections classified as group 3 were distinguished by a slower upslope, resulting from low average values of both viral production and infectivity (**Fig. S2k,l**). Group 4 infections had a rapid, high peak viral load due to high viral production and infectivity, as well as a relatively low average values for *ϕ*, the conversion to refractory state. Higher values of viral production and viral infectivity and low conversion to a refractory state mean that these infections rapidly burn through susceptible cells and target cell limitation slows the infection (**Fig. 4b, Fig.S2 j-l**). Only when the viral load was already decreasing did acquired immunity typically initiate to help clear the infection (**Fig. 4e)**. Infections in group 5 had a late, low peak and a long duration. Similar to group 3, the late peak was due to low rates of viral production and low infectivity (**Fig. S2k-l**). However, unlike group 3, these individuals also had low values of infected cell clearance, *δ*, and a very late onset of acquired immunity *τ*, allowing infection to persist (**Fig. S2n-o**). Finally, group 6 consisted of long infections with a high peak viral load. These infections were distinguished by high viral production and infectivity (**Fig. S2k-l)**, and globally weak immune responses including refractory cell conversion (**Fig. S2j)** and time-independent infected cell clearance rates (**Fig. S2n)**. Thus, late-acting acquired immunity was often required to clear the infection (**Fig. 4e and Fig. S2o,q)**, Overall, these results suggest that a complex interplay of viral and immune features dictate how individual infections differ according to peak viral load, viral expansion rate, viral clearance rate, and duration of shedding.

We next examined correlations between estimated model parameters and found several significant patterns (**Fig S3**). Viral infectivity, *β*, had a strong positive correlation with viral production rate, *π*. The viral production rate also had a positive correlation with the intensity of time-independent cytolytic immune pressure, *δ*. There was a strong negative correlation between the rate of reversion from refractory back to susceptible cells, *ρ*, and time of onset of late cytolytic immune pressure, *τ*. These results may suggest that viral fitness properties are related or that the durability of early innate responses is inversely correlated with the onset of delayed acquired immunity, but we cannot disentangle true biological correlations from potential identifiability challenges in the model structure.

### Lower peak viral load and earlier clearance during re-infection with omicron due to more effective early immune responses and more rapid late responses

The NBA cohort data set documented initial infection and reinfection in 67 individuals (**Fig. 5a, S4)**. Of the first infections, 52 were caused by a pre-delta variant and 15 by delta. For all individuals, the second infection was caused by an omicron variant. The mean peak viral load documented in the URT for a re-infection was 0.5 log_10_ viral RNA copies/ml lower than the mean for first infections. Though there was a slight negative correlation between peak viral load during the first infection and that of the second infection (**Fig. 5b**), the relationship was not statistically significant (*r_pearson_* = − 0.18, *p* = 0.15). The median time to clearance for reinfections was 7.5 days after detection compared with 11 days after detection for first infections (**Fig. 5c**). Using a slightly different data from the NBA cohort, Kissler et al. found evidence that an individual’s relative clearance speed is roughly preserved across infections^37^, prompting us to investigate whether this relationship, or others, appear in our model fits. We tested whether relative viral peak, time to peak, infection duration, or area under the curve were conserved by looking at the pearson correlation and did not find any significant relationships (p<0.05). We also checked whether estimated parameter values were conserved across sequential infections in the same individual, and again found no significant correlations across infections in the same individual.

**Figure 5:**
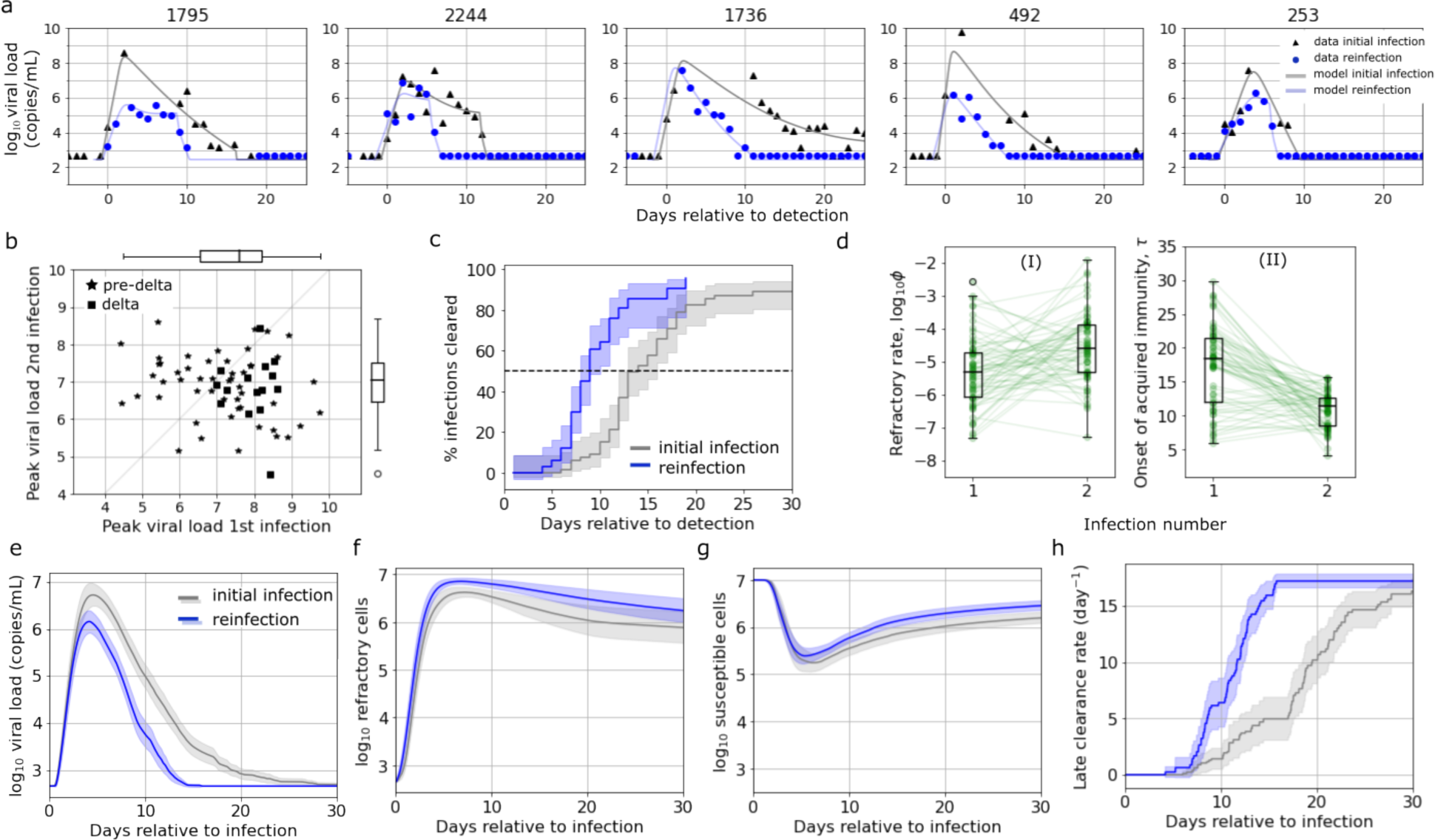
Mechanistic underpinning of more rapid clearance of SARS-CoV-2 during re-infection versus initial infection. Initial infection and re-infection were documented for 67 individuals in the NBA cohort. (a) Examples of data and model fits for infection and reinfection in the same individual (b) As measured from the data, peak viral load of reinfection against peak viral load of first infection. In all cases the variant causing the reinfection was omicron, and the variant causing the first infection was either delta or a pre-delta variant. The mean peak viral load was around 0.5 log lower for second infection (t-test statistic = 2.26, p = .0254) (c) Proportion of infections cleared for reinfection (blue) and first infections (gray) over time, as measured from the data. Median time to clearance is 7.5 vs. 12 days since detection. (d) Boxplots of estimated individual parameters for infection and reinfection that are significantly different between the two groups (p_adjusted_ < 0.05 for Mann-Whitney U-test). During re-infection with omicron, the rate that susceptible cells convert to a refractory state is higher and the onset of the late immune response occurs significantly earlier. (e) Mean viral load, (f) number of refractory cells, (g) number of susceptible cells, and (h) late clearance rates over time for the two groups as predicted by mechanistic model.

Two model parameter values were significantly different between first versus second infections (**Fig. 5d**). Reinfections had higher values of *ϕ*, indicating a faster conversion of susceptible cells to a refractory state. This more potent early immune responses contributed to lower peak viral loads. The timing of the acquired immune response, *τ*, was also earlier, during reinfection suggesting more rapid activation of immune memory. We found that parameter values estimated during first infection were not predictive of parameter values estimated for a sequential infection in the same individual. Mean model projections recapitulated viral load patterns observed in the data (**Fig 5e**). We plotted cell populations from the model simulations for the two groups, and reinfection appeared to result in more refractory cells (**Fig 5f**) and a smaller decrease in susceptible cells **(Fig 5g)**. The acquired immune response initiated sooner and at a higher magnitude during re-infection (**Fig 5h**).

### Waning early immune response and strong initial clearance of infected cells as a cause of off-therapy viral rebound

Recent studies have shown that viral rebound during the natural course of untreated SARS-CoV-2 infection is relatively common, occurring in over 10% of cases by some estimates^38^ (https://www.fda.gov/media/166197/download), though rates vary according to definition. In their analysis of the NBA cohort, Hay et al. flagged 40 out of 1334 cases (3%) as rebound, defined by a non-monotonic sequence of test results^36^. As their most inclusive definition of rebound, they identified cases that achieved an initial clearance of at least 2 days with cycle threshold greater than or equal to 30, followed by at least 2 days with cycle threshold < 30.

We defined simulated infections as rebound if there were 2 or more peaks with height > 3 log_10_ RNA copies/ml and prominence > 0.5 log_10_ RNA copies/ml. We defined prominence as the height above the preceding local minima, as illustrated in (**Fig. 6a**). With these criteria, we identified 7.0% of the 1510 cases as rebound. These cases are marked with an “R” and included first in **Fig S6**. Note that we were unable to connect viral rebound to recrudescence of COVID-19 symptoms because we do not have daily reports of symptom status.

**Figure 6:**
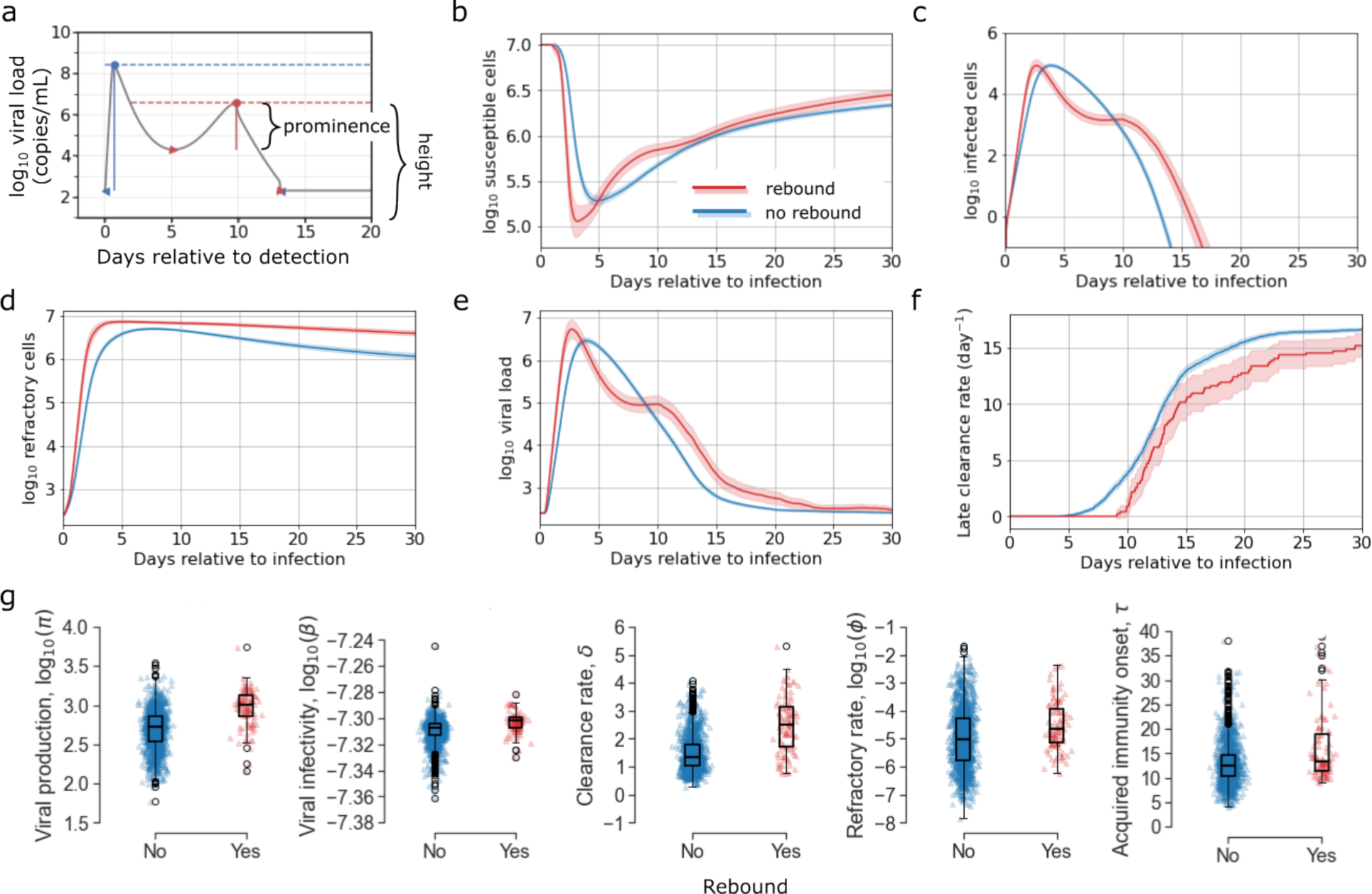
Model fiPng to viral rebound in the NBA cohort. (a) We classified infections as examples of viral rebound if there are at least two peaks in the model simulation with height of 3 logs and prominence of 0.5 log. Mean (b) number of susceptible cells, (c) number of active infected cells, (d) number of target cells that are refractory, (e) viral load, and (f) rate of late clearance as predicted by our mathematical model for rebound vs. non-rebound cases in red and blue respectively. 95% confidence interval shaded. (g) Distribution of individual parameter estimates for the rebound vs. non-rebound cases. Only those for which the mean differs significantly are displayed (p_adjusted_ < .05 for Mann-Whitney U test).

We observed several differences between rebound and non-rebound infection. In cases of rebound, susceptible cells were lost more rapidly initially but also replenished earlier from the refractory compartment (**Fig. 6b)**. On average, rebound trajectories had both an earlier peak in infected cells and an earlier, higher peak in refractory cells (**Fig. 6c-d**). The large number of refractory cells drives a more rapid replenishment of susceptible cells. The persistence of infected cells with re-emrgence of susceptible cells allowed for a second surge of viral production, which had been reduced by fast early clearance of infected cells (**Fig 6e**). The delayed onset of the late acquired immune response also allowed sufficient time for this to occur before the infection was ultimately cleared (**Fig 6e-f**). Cases with rebound had higher viral production rates, *π*, and higher viral infectivity, *β*, which combined to allow for growth of the viral population even with a reduced number of target cells. Rebound cases also had a higher clearance rate, *δ*, and a faster conversion of susceptible cells to a refractory state. Together, these forces preserve susceptible cells through the rapid initial clearance of infected cells and protection in a refractory state. Notably, rebound cases did not have a significantly higher reversion rate, *ρ*, to account for replenishment of susceptible cells after the first viral peak. Rather, the faster replenishment of susceptible cells occurs due to the high number of refractory cells. The viral rebound group also had a delayed onset of late immune killing, *τ*, which allowed time for two peaks to occur before pressure from the acquired immune system cleared the infection (**Fig 6g**).

## Discussion

Viral kinetics are vital to understanding the pathogenesis of infection and, ultimately, to optimizing therapies. Here, we use a remarkable cohort from the NBA, which is unique both for its size and because it captures early pre-symptomatic timepoints during infection, to describe the increasing variability in viral load patterns observed in SARS-CoV-2 infected people. We observe that with a general increase in population level immunity due to prior infection and vaccination, peak viral load is often lower and earlier with more rapid elimination of virus.

Our mathematical model identifies testable mechanistic hypotheses for these observed differences. We first predict that low peak viral loads are associated with lower viral production within infected cells and lower viral infectivity. Moreover, for viral loads that also peak early (observed in group 1), the model predicts a rapid conversion of susceptible cells to a refractory state. Both effects are compatible with data observed in animal models and *in vitro* models describing effects of interferon which limit the extent of viral replication and protect uninfected cells from viral entry^39–43^. Appropriate follow up experiments to validate this prediction would include local sampling of nasal cytokines and other mediators of local immunity during critical early timepoints of infection as has been done in humans for other respiratory viral infections^44^.

Different viral shedding patterns are also driven by varying balances between the magnitude of the early cytolytic immune response, which wanes as the number of infected cells and viral load declines following peak, and the late sustained immune response. In our model, we assumed this response does not dissipate with decrease in virus, so we hypothesize that most of the late response is acquired and due to either expanding T cell or antibody levels. Prior work suggested that during primary infection, plasma SARS-CoV-2 IgG levels rise too late to explain reduction in viral load^45^. However, this study was performed in an immunologically naïve cohort and needs to be reassessed in the current infection environment^46, 47^. T cell mediated killing of infected cells may also assist in elimination of infected cells during infection^46, 48^. The results from our study suggest that at the time of the NBA cohort, substantial differences in timing and intensity of acquired immune responses were still present and contributed to variability in viral kinetics.

Our results suggest that upon reinfection, the early/innate response and the late acquired response are both more effective. The mechanisms underlying this observation are unclear. One possibility is that a higher density of tissue resident NK cells, B cells and T cells may exist after first infection and vaccination. In other viral infections, it has been observed that an increase in pre-infection tissue resident T cells predicts earlier initiation of a local innate and acquired response due to early antigen recognition^49, 50^. These model predictions merit experimental follow up.

Unfortunately, we are not able to link the heterogenous virologic patterns observed in the NBA cohort with severity of symptoms or future development of post-acute sequelae of SARS-CoV-2 infection as this data was not available. For multiple other viruses, viral loads have been identified as relevant correlates of disease^51–54^, and late SARS-CoV-2 viral loads have been linked with severity of infection among hospitalized people^55, 56^. During clinical trials, reductions in nasal viral load due to monoclonal antibodies, nirmatrelvir / ritonavir, and molnupiravir correlated with very large reductions in the incidence of hospitalization and death^28, 29^. Yet, early remdesivir which had a large clinical benefit was associated with no viral reduction in nasal passage several days after treatment^30^, highlighting that key viral load surrogates may be in the lung rather than nasal passages or that early viral loads are more predictive of outcome^22^. Because early and peak viral load measurements are so rarely obtained during COVID-19 infection, the clinical importance of these values remain unknown.

Several further limitations of this work are important to highlight. An issue that is universal to the field is that our model does not capture anatomic compartmentalization of viral shedding. Our previous model demonstrated in non-human primates that SARS-CoV-2 kinetics in the lung differ in subtle but important ways from those in the upper airways, and that these differences are particularly significant in the context of antiviral therapy^22^. It is likely that our subgroups of shedding may cluster differently if we had access to serial whole lung viral loads. The re-seeding of infection in the nose from the lungs or vice versa may also provide alternative explanations for the dynamics observed in this data set, particularly viral rebound. Unfortunately, such detailed studies are not available in any human cohort. Studies using saliva do suggest slightly different kinetics than those from nasal swabs^57^, but it is doubtful that saliva captures total viral load in the lung.

Another issue shared by all mathematical models in the field is the lack of sufficiently granular, tissue-based immune data to precisely model the innate and acquired immune responses. Rather, our model uses several terms to capture the timing and intensity of what is likely to be a complex, multi-component response. As with multiple other respiratory virus models and based on experimental data showing that interferon-alpha protects cells from infection, we assume that infection temporarily makes susceptible cells refractory to viral entry^20–22, 40, 43^. Finally, we assume a late, sustained immune response that varies by intensity and timing, compatible with an acquired memory response.

A final limitation shared by all intra-host SARS-CoV-2 models in humans is that we are not able to measure potentially important initial conditions of infection, including viral inoculum and the number of immune cells within a relevant spatial microenvironment of infection. Thus, the model may over ascribe observed differences in observed viral load trajectories to differences in immune responses rather than exposure viral load.

In summary, we identify distinct shedding patterns in adults with SARS-CoV-2 infection, with shorter, lower viral load infection more commonly observed in persons with omicron infection, prior vaccination, and recent prior infection. The mechanistic predictors of rapidly contained infection are more rapid conversion of susceptible cells to a refractory state along with more rapid and intense late cytolytic immune responses.

## Materials and Methods

### Study Overview

We analyzed SARS-CoV-2 viral load data collected during untreated infections in the NBA cohort. We clustered this data into 6 dynamic groups, which were statistically different in terms of peak viral load, time to peak viral load, area under the viral load curve, and time to clearance. Drawing on previous models in the field, we developed a set of candidate ordinary differential equation (ODE) mathematical models. We then used model selection theory to determine which version the data supported most strongly. With a validated model of SARS-CoV-2 infection, we examined which parameter values differ to explain the variable viral shedding patterns observed in the six dynamic groups. We also used this approach to explain the differing dynamics of first and second infections captured in the NBA cohort, and to explain the mechanisms underlying viral rebound.

### Data Pre-processing

We used data from the NBA cohort previously published by Hay et al.^36^. The group documented 2875 individual SARS-CoV-2 infections in 2678 people through frequent quantitative PCR testing. First, we filtered this data to include only infections with at least 4 positive quantitative samples to provide adequate viral dynamics data for model fitting. This yielded 1510 infections in 1442 individuals, of which 177 were caused by a pre-VOC variant, 46 by alpha, 163 by delta, and 1124 by omicron (**Fig. 1a**). We further identified a “well-documented” subset of these infections by filtering for infections that had their first quantitative test within 5 days of detection and included test results through 20 days after detection or confirmed elimination of virus prior to day 20 (two consecutive negative tests). This well-documented group consisted of 810 individual infections in 768 people. For clustering, we randomly chose one infection to retain from each individual with multiple documented infections, resulting in a group of 768 infections in 768 individuals. We also filtered the well-documented group for infections with a negative test result within 2 days prior to detection, yielding 266 cases with both early detection and 3 weeks of documentation. We refer to this subset as “fully documented.”

### Quantitative Features of Viral Dynamics

To convert cycle threshold (Ct) values to viral genome equivalents, we averaged Ct1 and Ct2 for each individual and applied equation S2 from Kissler et al.^58^ That is,

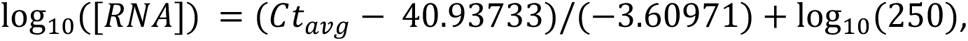

where the concentration of viral RNA is in copies/ml.

We calculated the peak viral load for a given infection as the maximum measured log_10_ viral load over all quantitative data points and the time to peak viral load was the day of this measurement. We calculated the area under the log_10_ viral load curve from the date of detection through the last quantitative measure of viral load, linearly imputing missing values between data points. Note that this quantity is an underestimate for individuals without confirmed clearance. We calculated the median time to clearance by identifying when the cumulative incidence curve for clearance of the virus crossed 50%. The cumulative incidence curve is the inverse of the Kaplan-Meier curve for survival of the virus. The Kaplan-Meier curve, KM, and confidence interval was computed using the Python package scikit-survival 0.21.0 (https://scikit-survival.readthedocs.io/en/stable/). The cumulative incidence curve is then 1-KM.

### Data Clustering

We clustered well-documented infections into 6 dynamic groups using k-means clustering as implemented in the Python package scikit-learn 1.2.2 (https://scikit-learn.org/stable/modules/generated/sklearn.cluster.KMeans.html). As input features, we used these 21 daily test results. These came from the day infection was detected through 20 days after detection. If any daily measurements were missing between recorded test values, we imputed the missing measurements linearly. If the last test date for an individual was prior to day 20, so there were missing daily measurements after the last test, we appended negative test values to reach 20 days (**Fig S5a**). This occurred only for infections for which clearance was confirmed with 2 consecutive negative tests, since we clustered well-documented infections.

To select these hyperparameters for the k-means clustering, we tested values of k from 2 to 20 for three possible interpolation methods, linear, quadratic, or cubic spline, and two possible surveillance periods, 13 or 20 days post detection (2 or 3 weeks surveillance). Comparing these scenarios, linear interpolation up to 20 days post detection had the lowest within cluster sum of squares (**Fig. S5b**). Based on the location of the “elbow” in the plots, we chose to proceed with k = 6 clusters. Using k < 6 results in less distinctive behaviors between the groups, while using more clusters resulted in some non-interpretable cluster centers (**Fig. S5c-g).**

### Mathematical Model of SARS-CoV-2 Dynamics

We considered several ordinary differential equations models for SARS-CoV-2 infection dynamics. The full model tracks the number of target cells that are susceptible to infection (*S*), target cells that are refractory to infection (*R*), infected cells in an eclipse phase (*I_E_*), infected cells actively producing virus (*I_P_*), and SARS-CoV-2 virions (*V*). Susceptible cells are infected at rate *βSV*, and become refractory at rate *ϕI_p_S*. Refractory cells revert to a susceptible state at rate *ρR*. When cells are first infected, they enter an eclipse phase, from which they transition to a state of producing virus at rate *k*. Productively infected cells are cleared at rate 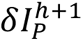, where the dependence on infected cells reflects an innate immune response with no memory. When the duration of infection surpasses time *τ*, the clearance rate of infected cells increases by *mI_P_*, capturing the delayed onset of a cytolytic acquired immune response with memory. Productively infected cells produce virus at rate *π*, and free virions are cleared at rate *γV*. Under these assumptions, the model has the form:

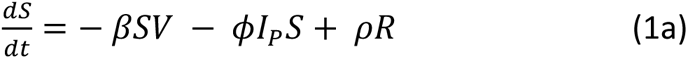

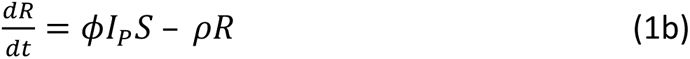

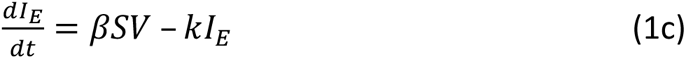

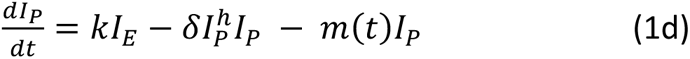

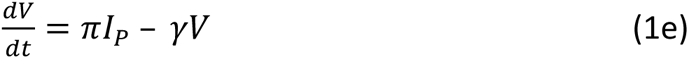

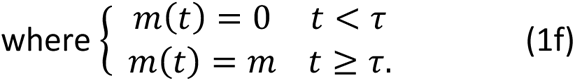

To ensure that the model did not predict spurious oscillations in viral dynamics, we enforced that viral production was zero when *I_P_* was less than 1. In the optimal model, the parameter h = 0 for all individuals, so the early per-cell clearance rate of infected cells is not density dependent.

As initial conditions, we set (*S*_0_, *R*_0_, *I_E_*_0_, *I_P_*_0_, *V*_0_) = (1 × 10^7^, 0, 0, 0, *V*_0_). Previous models of SARS-CoV-2 infection in the nasal compartment have used an initial value of 10^7^ − 10^8^ susceptible cells, based on estimates that 2-20% of epithelial cells in the upper respiratory tract display the ACE 2 receptor^59–61^. We assumed that the initial number of refractory cells is zero, because the early immune response is inactive prior to infection. We initiated simulations with zero infected cells, so *I_E_*_0_= *I_P_*_0_ = 0, and a small viral inoculum to reflect the tight bottleneck that transmission places on viral replication. The number of virions present at the outset of infection was assumed to be below the limit of detection, but the precise inoculum was initially allowed to vary for individuals. During model fitting, we estimated the onset of infection relative to detection (date of first positive test), noting this difference as *t*_0_. Among individuals in the NBA cohort for whom symptom onset was known, the mean time of symptom onset was the date of detection, so *t*_0_ is correlated with the incubation period of SARS-CoV-2. With this in mind, we restricted estimates of *t*_0_ to fall between 0 and 20 days based on a 2022 review by Wu et al., which reported that across 142 studies of SARS-CoV-2 infection, the incubation period ranged from 1.80 to 18.87 days^62^.

To maintain identifiability, we fixed two parameter values, setting the rate of viral production onset to be *k* = 4 in accordance with Ke at al.^23^ and the rate of clearance of free virions to be *γ* = 15 in accordance with Goyal et al.^11^

### Model Fitting and Selection

We fit the model in Eq. 1, as well as simpler versions that eliminate one or more immune components and/or the eclipse phase, to data from the NBA cohort using a non-linear mixed effect approach^63^. With this approach, a viral load measurement from individual *i* at time point *k* is modeled as log_10_(*y_ik_*) = *f_V_*(*t_ik_, θ_i_*) + *ϵ*, where *f_V_* represents the solution of the ODE model for the state variable describing the virus, *θ_i_* is the parameter vector for individual *i*, and *ϵ* ∼ *N*(0, *σ*^2^) is the measurement error for the log_10_-transformed viral load data. Furthermore, in the population model, each individual’s parameters can be written as the sum of the average population value, *θ_pop_*, and a random effect encompassing their deviation from the average, *η_i_*. That is, the parameters for individual i are given by *θ_i_* = *θ_pop_* + *η_i_*. We fixed *σ* = 0.5 log_10_ viral RNA copies/ml when comparing model fits, so that any differences in likelihood of the full model occur due to a change in agreement between model simulations and data rather than a drastic increase in the estimated magnitude of the measurement error.

For model selection, we first worked with the 266 fully documented infections (early detection and at least 3 weeks of follow-up or confirmed clearance). In addition to the raw data, for individuals without confirmed elimination we imputed 5 “assumed negative” test results at 2-day intervals starting at 40 days post-detection. Out of the 1510 infections considered in model fitting, 629 had regular measurements past 40 days and 99.5% of tests collected past day 40 were negative. For viral load observations below the lower limit of quantification or marked as “assumed negative”, we used the probabilistic model that Monolix software provides for left-censored data (https://monolix.lixoft.com/censoreddata/).

The candidate models that we considered are listed in the supplementary material (**Table S1**). For each candidate model, we used the Stochastic Approximation of the Expectation Maximization (SAEM) algorithm embedded in the Monolix software to obtain the Maximum Likelihood Estimation (MLE) of the vector of fixed effects, *θ_pop_*, and the MLE of the vector of standard deviations of the random effects, *σ*_*θ*_, for the model parameters *β*, *π*, *ϕ*, *ρ*, *δ*, ℎ, *τ*, *m*, the delay between infection and date of detection, *t*_0_, and the initial viral inoculum, *V*_0_ (https://monolix.lixoft.com/tasks/population-parameter-estimation-using-saem/). We assumed a lognormal distribution for parameter values, and a logit distribution for initial conditions. The delay between infection and detection, *t*_0_, was assumed to fall between 0 and 20 days. The viral inoculum was assumed to fall between 0 and 250 log_10_ viral RNA copies/ml.

We ran the SAEM algorithm six times for each model using randomly selected initial values for the estimated parameters. Using the parameter set with the highest likelihood, we computed the Akaike Information Criterion (AIC) for each model. Recall that *AIC* = −2 max(log ℒ) + 2*m* where ℒ is the likelihood that the data was generated by this model with these parameter values and *m* is the number of model parameters. Hence smaller AIC scores indicate that a model is statistically more likely to explain the data. The model with the smallest AIC score in the initial model selection phase included an eclipse phase, a refractory cell compartment and time-dependent clearance of infected cells, but not density dependent clearance. All AIC scores are recorded in **Table S1**.

The best fit run for the optimal model estimated very little variation in the viral inoculum between individuals. The population average was, *V*_0_*pop*_ = 97.3 while the standard deviation of the distribution of random effects was only 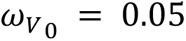, suggesting that fixing this parameter at the same value for all individuals may still allow for reasonable fits. We fixed *V*_0_near the estimated population mean, *V*_0_ = 97 for all individuals and re-ran the SAEM algorithm. This yielded very similar fits to the best fit from Table S1 with a slightly lower AIC score of 13731 compared to 13738. While we expect that the actual viral exposure initiating individual infections in the NBA cohort varied, this suggests that estimating *V*_0_, *π*, and *t*_0_ simulataneously for each individual does not lend additional flexibility. For further model fitting, we kept *V*_0_ fixed at 97.

To test whether variability in viral dynamics can be attributed to differences in prior exposure, age, or infecting lineage, we performed one-way ANOVA for the random effects of each of the estimated model parameters against these covariates (implemented in monolix). In this case, the null-hypothesis is that the mean of the random effects (calculated from the individual parameters sampled from the conditional distribution) is the same for each category of the categorical covariate. Ranking all possible covariates by their p-value, the most likely covariate was between the onset of acquired immunity and vaccination status. We tried adding this as a covariate to the model, which allows for a perturbation of the population mean, *τ_pop_*, by some value *β*_τ__**j** for each possible vaccination status j. Including this covariate improved the model fit according to AIC score, improving from 13731 to 13627. We next checked whether this was a meaningful addition to the model with the Wald test, which tests the null hypothesis that *β*_τ__*j* = 0 for each possible vaccination status j. Infections in unvaccinated individuals were significantly different from infections in individuals who had been boosted (*β*_τ_ _0 doses = 0.73, p <2.2e-16) and the group that had no record(*β*_τ_ _ no record = 0.44, p=6.77e-7). However, individuals who had received 1, 2 or 3+ vaccinations were not significantly different from each other (*β*_τ_ _ 1dose = -.36, p=3.6e-1; *β*_τ_ _ 2 dose = -.076, p=3.18e-1). This prompted us to regroup vaccination status into a new categorical covariate, indicating unvaccinated, at least one dose, or no record. With this model, the onset of acquired immunity differed significantly for infections in unvaccinated individuals vs. those who received at least one dose of the vaccine (*β*_τ_ _ >1dose = -0.8, p<2.2e-16), but the difference between infections in unvaccinated individuals and those with no record was not significant (*β*_τ_ _ no record= -0.2, p = 2.03e-2). Then we regrouped vaccination status into just two categories, one being individuals who are unvaccinated or have no record and the second being individuals with a record of 1 or more SARS-CoV-2 vaccinations. We repeated this process of choosing one new covariate to add according to the lowest significant p-value resulting from the ANOVA, testing its utility using the AIC and Wald test, and coarsening the categorization if indicated, until no further significant p-values resulted from the ANOVA. This resulted in three covariates, unvaccinated/ no record versus at least one recorded vaccination modified the onset of acquired immunity *τ* and the infecting lineage being pre-VOC/delta versus omicron versus unknown modified both the onset of acquired immunity *τ* and the rate at which susceptible cells enter a refractory state, *ϕ*. Including these covariates reduced the AIC score by 149 to 13589. The models tested along the way are reported in **Table S2**.

For the best fitting model, there were significant correlations between the random effects of model parameters *β*, *π* and *δ*, as well as *ρ* and *τ*. We started with the best model from Table S2 and allowed for linear correlations between these parameters in the final model (https://monolix.lixoft.com/statistical-model/individual-model/individualdistribution/). This further improved the AIC score by 111 points (**Table S3**). The correlation structure of the final set of parameter estimates is shown in **Figure S3** and the correlations between the random effects can be found on the github page at https://github.com/lacyk3/SARS-CoV-2Kinetics. Once the final model was selected, we further restricted the standard deviation of the measurement error to *σ* = 0.4 log_10_ viral RNA copies/ml to capture examples of viral reboundin the data and ran the SAEM algorithm in Monolix to estimate parameters for all 1510 infections. Population parameter values are included in **Table S4** and individual model fits are shown in **Figure S6**. Estimated individual parameter values are accessible at https://github.com/lacyk3/SARS-CoV-2Kinetics.

## Statistics

When comparing quantitative features and parameter values across different groups, we used a two-sided Mann-Whitney U-test. When assessing significance of the results, we adjusted p values using the Bonferroni correction for the number of comparisons before comparing against a significance threshold of p > 0.05.

## Data and Code availability

The data analyzed in this work was previously published by Hay et al. and is available on github at https://github.com/gradlab/SC2-kinetics-immune-history. The code for generating all analysis and figures included in this manuscript is available at https://github.com/lacyk3/SARS-CoV-2Kinetics.

## Acknowledgements

We thank Yonatan Grad, Stephen Kissler, and the members of the NBA cohort for making this data available. We also thank Dan Reeves and Liz Duke for their helpful advice regarding figures. This work was supported by National Institutes of Health (NIH) grants R01AI169427 & R01AI121129.

## Author contributions

K.O. and J.T.S. conceptualized the study and developed mathematical models. K.O. performed experiments and statistical analysis. K.O., S.E., and J.T.S. interpreted results and wrote the manuscript.

