## Supplementary Materials for "Heterogeneous SARS-CoV-2 kinetics due to variable timing and intensity of immune responses"

***Table S1: Selecting the best model for fit to longitudinal viral load data by comparing AIC score.*** *Initial conditions for all models were* $S_0 = 1 \times 10^7, I_0 = 0, R_0 = 0,$ and $E_0 = 0$. *The initial viral inoculum was estimated from a logit distribution with lower bound of 0 and upper bound of 250 viral RNA copies/ml. The delay between infection and detection was estimated from a logit distribution with lower bound of 0 and upper bound of 20 days. The standard deviation of measurement error was fixed at a = 0.5* $\log_{10}$ *copies viral RNA/mL for all runs to ensure that variation in AIC scores came from differences in the ODE models. A subset of the full NBA data consisting of 266 well-documented infections was used for model comparison. An approximation of the AIC score was computed for each model by running the SAEM algorithm from six different initial parameter guesses and then taking the lowest score, indicated with bold and underline for each model. The best model overall was M7.*

| Model | AIC |
| --- | --- |
| ***M1:*** Target-cell limited model<br>$\frac{dS}{dt} = -\beta SV$ (S1a)<br>$\frac{dI_P}{dt} = \beta SV - \delta I_P$ (S1b)<br>$\frac{dV}{dt} = \pi I_P - \gamma V$ (S1c) | 16102, **<u>16032</u>**, 16063, 16069, 16042, 16049 |
| ***M2:*** Target cell limited model with eclipse phase (TEIV)<br>$\frac{dS}{dt} = -\beta SV$ (S2a)<br>$\frac{dI_E}{dt} = \beta SV - kI_E$ (S2b)<br>$\frac{dI_P}{dt} = kI_E - \delta I_P$ (S2c)<br>$\frac{dV}{dt} = \pi I_P - \gamma V$ (S2d) | 15716, 15706, 15697, **<u>15647,</u>** 15698, 15685 |
| ***M3:*** TEIV model with time-dependent clearance of infected cells<br>$\frac{dS}{dt} = -\beta SV$ (S3a)<br>$\frac{dI_E}{dt} = \beta SV - kI_E$ (S3b)<br>$\frac{dI_P}{dt} = kI_E - m(t)I_P$ (S3c)<br>$\frac{dV}{dt} = \pi I_P - \gamma V$ (S3d)<br>where $\begin{cases} m(t) = 0 & t < \tau \\ m(t) = m & t \geq \tau. \end{cases}$ | 14823, 14792, 14846, 14799, **<u>14782</u>**, 14799 |

| | |
|---|---|
| ***M4:*** TEIV model with refractory cell compartment (TREIV)<br>$\frac{dS}{dt} = -\beta SV - \phi I_P S + \rho R$ (S4a)<br>$\frac{dR}{dt} = \phi I_P S - \rho R$ (S4b)<br>$\frac{dI_E}{dt} = \beta SV - kI_E$ (S4c)<br>$\frac{dI_P}{dt} = kI_E - \delta I_p$ (S4d)<br>$\frac{dV}{dt} = \pi I_P - \gamma V$ (S4e) | 14865,<br>14879,<br>**14861**,<br>14888,<br>14879,<br>14862 |
| ***M5:*** TIEV model with density dependent clearance of infected cells<br>$\frac{dS}{dt} = -\beta SV$ (S5a)<br>$\frac{dI_E}{dt} = \beta SV - kI_E$ (S5b)<br>$\frac{dI_P}{dt} = kI_E - \delta I_P^h I_P$ (S5c)<br>$\frac{dV}{dt} = \pi I_P - \gamma V$ (S5d) | 15680,<br>15652,<br>15736,<br>**15629,**<br>15730,<br>15669 |
| ***M6:*** TEIV model with time and density-dependent clearance of infected cells<br>$\frac{dS}{dt} = -\beta SV$ (S6a)<br>$\frac{dI_E}{dt} = \beta SV - kI_E$ (S6b)<br>$\frac{dI_P}{dt} = kI_E - -\delta I_P^h I_P - m(t) I_P$ (S6c)<br>$\frac{dV}{dt} = \pi I_P - \gamma V$ (S6d)<br>where $\begin{cases} m(t) = 0 & t < \tau \\ m(t) = m & t \geq \tau. \end{cases}$ | 14376,<br>**14360,**<br>14433,<br>14589,<br>14534,<br>14570 |
| ***M7:*** TREIV model with time-dependent clearance of infected cells<br>$\frac{dS}{dt} = -\beta SV - \phi I_P S + \rho R$ (S7a)<br>$\frac{dR}{dt} = \phi I_P S - \rho R$ (S7b) | 13748,<br>13785,<br>13775,<br>13776,<br>13778,<br>**13738*** |

| | |
|---|---|
| $\frac{dI_E}{dt} = \beta SV - kI_E$ (S7c)<br><br>$\frac{dI_P}{dt} = \mathrm{kI_E} - \delta \mathrm{I_P} - \mathrm{m}(t)I_P$ (S7d)<br><br>$\frac{dV}{dt} = \pi I_P - \gamma \mathrm{V}$ (S7e)<br><br>where $\begin{cases} m(t) = 0 & t < \tau \\ m(t) = m & t \geq \tau. \end{cases}$ | |
| ***M8:*** TREIV model with density-dependent clearance of infected cells<br>$\frac{dS}{dt} = -\beta SV - \phi I_P S + \rho R$ (S8a)<br><br>$\frac{dR}{dt} = \phi I_P S - \rho R$ (S8b)<br><br>$\frac{dI_E}{dt} = \beta SV - kI_E$ (S8c)<br><br>$\frac{dI_P}{dt} = kI_E - \delta I_P^h I_P$ (S8d)<br><br>$\frac{dV}{dt} = \pi I_P - \gamma V$ (S8e) | **<u>14916,</u>**<br>15166,<br>15225,<br>15361,<br>15373,<br>15077 |
| ***M9:*** TREIV model with time and density-dependent clearance of infected cells<br>$\frac{dS}{dt} = -\beta SV - \phi I_P S + \rho R$ (S9a)<br><br>$\frac{dR}{dt} = \phi I_P S - \rho R$ (S9b)<br><br>$\frac{dI_E}{dt} = \beta SV - kI_E$ (S9c)<br><br>$\frac{dI_P}{dt} = kI_E - \delta I_P^h I_P - m(t)I_P$ (S9d)<br><br>$\frac{dV}{dt} = \pi I_P - \gamma V$ (S9e)<br><br>where $\begin{cases} m(t) = 0 & t < \tau \\ m(t) = m & t \geq \tau. \end{cases}$ (S9f) | **<u>13797,</u>**<br>13827,<br>13800,<br>13837,<br>13841,<br>13821 |

***Table S2: Incorporating covariates.*** *To further improve model fits, we considered including covariates accounting for the strain of the infection, age group, the vaccination status or cumulative prior exposures status at the time of infection. The fitting conditions were the same as described in Table S1, and the model used was M7. The best set of co-variates included no dose / no record versus at least one vaccine dose, as well all non-omicron variants versus omicron variants.*

| Covariates | AIC scores across 5 runs | Notes |
|---|---|---|
| vaccination status, $\tau$ | 13710,<br>**13627,**<br>13787,<br>13849,<br>13794 | Wald test $\tau$ – difference from boosted<br>- 0 dose: 0.73, p < 2.2e-16<br>- *1 dose: -0.36, p = 3.6e-1*<br>- *2 dose: -0.076, p = 3.18e-1*<br>- No record: 0.44, p = 6.77e-7 |
| 0 dose v. >1 dose v. no record, $\tau$ | 13700,<br>13677,<br>**13665**,<br>13713,<br>13690 | Wald test $\tau$ – difference from 0 doses<br>- >1 doses: -0.8, p < 2.2e-16<br>- *No record: -0.2, p = 2.03e-2*<br>Next most likely covariate suggested is $\phi$ for lineage (ANOVA: 13.92 , p = 1.89e-8 ) |
| 0 dose/no record v. >1 dose, $\tau$ ;<br>lineage, $\phi$ | 13652,<br>**13608**,<br>13658,<br>13665,<br>13634 | Wald test $\tau$ – difference from 0 doses/no record<br>- 1+ doses: -0.7, p = 3.61e-5<br>Wald test $\phi$ – difference from delta<br>- *Other: 0.46, p = 2.04e-1*<br>- Omicron: 1.5, p = 2.72e-5<br>- Unknown: 2.23, p = 2.92e-6 |
| 0 v. >1 dose, $\tau$ ;<br>other/delta v. omicron, $\phi$ | 13666,<br>**13637,**<br>13649,<br>13673,<br>13647 | Wald test $\phi$ —difference from delta/other<br>- Omicron: 1.08, p = 1.02e-9<br>- Unknown: 1.88, p = 2.31e-7<br>Next most likely covariate suggested is $\tau$ for other/delta v. omicron v. unknown (ANOVA: 6.23, p = 2.26e-3) |
| 0 dose/no record v. >1 dose, $\tau$ ;<br>other/delta v. omicron, $\tau, \phi$ | 13616,<br>**13589*,**<br>13601,<br>13619,<br>13593 | Wald test—difference from delta/other<br>- Omicron: -.54, p = 1.92e-12<br>- Unknown: -.6, p = 8.91e-7<br>Next most likely covariate suggested is $m$ for 0 v. >1 dose (ANOVA: 3.21 , p = 7.42e-2) |
| 0 dose/no record v. >1 dose, $\tau, m$ ;<br>other/delta v. omicron, $\tau, \phi$ | **13596,**<br>13603,<br>13617,<br>13610,<br>13633 | Wald test—difference from 0 doses/no record<br>- *1+ doses: 0.484, p = 8.8e-2* |

***Table S3: Incorporating correlations.*** *To further improve model fits, we used the best model from Table S2 and incorporated linear dependencies between correlated model parameters into the statistical model for the random effects. The Pearson correlation between the random effects for each pair of parameters was calculated, and the benefit of accounting for the most prominent correlations at the population level was tested by selecting the correlation groups indicated in parenthesis in column 2 in the monolix interface, re-running the SAEM algorithm five times, and comparing the resulting AIC scores.*

| Covariates | Correlated parameters | AIC | | | | |
|---|---|---|---|---|---|---|
| | | Run 1 | Run 2 | Run 3 | Run 4 | Run 5 |
| 0 v. >1 dose, $\tau$ ; other/delta v. omicron, $\tau, \phi$ | none | 13616 | **13589** | 13601 | 13619 | 13593 |
| 0 v. >1 dose, $\tau$ ; other/delta v. omicron, $\tau, \phi$ | $(\pi, \phi, \delta)$, $(\tau, \rho)$ | 13504 | 13499 | **13478*** | 13531 | 13541 |
| 0 v. > 1 dose, $\tau$ ; other/delta v. omicron, $\tau, \phi$ | $(\pi, \phi, \delta, \tau, \rho)$ | 13566 | 13540 | 13570 | **13536** | 13540 |
| 0 v. >1 dose, $\tau$ ; other/delta v. omicron, $\tau, \phi$ | $(\pi, \phi, \delta)$ | 13553 | 13559 | **13531** | 13565 | 13564 |
| 0 v. >1 dose, $\tau$ ; other/delta v. omicron, $\tau, \phi$ | $(\tau, \rho)$ | 13556 | 13585 | **13538** | 13564 | 13568 |

***Table S4: Population parameters from final model fits.*** *Using M7 with fixed initial conditions* $S_0 = 1 \times 10^7, I_0 = 0, R_0 = 0,$ and $E_0 = 0$, *and* $V_0 = 97$ *viral RNA copies/mL, and estimating the delay between infection and detection from a logit distribution with lower bound of 0 and upper bound of 20 days, we estimated model parameters to describe 1510 infections in the NBA cohort. We further included correlations between model parameters* $\pi, \phi, \delta$, *and* $\tau$, $\rho$. *For the mixed-effect model, the standard deviation of measurement error was fixed at a = 0.4* $log_{10}$ *copies viral RNA/ml. The population parameters are recorded here and estimated individual parameters are available at [https://github.com/lacyk3/SARS-CoV-2Kinetics](https://github.com/lacyk3/SARS-CoV-2Kinetics). The corresponding model fits are included in Figure S7.*

| Parameter (unit) | Symbol | Population Mean | Standard Error | Standard deviation of random effects | Distribution Of random effects | Source |
|---|---|---|---|---|---|---|
| viral infectivity ($\log_{10}$ (RNAcopies/mL)$^{-1}$ day$^{-1}$) | $\log_{10}\beta$ | -7.31 | 3.17e-3 | 4.69e-2 | normal | estimated |
| viral production rate ($\log_{10}$ day$^{-1}$ ) | $\log_{10}\pi$ | 2.74 | 9.7e-3 | 0.316 | normal | estimated |
| rate at which refractory cells revert to susceptible state ($\log_{10}$ day$^{-1}$ ) | $\log_{10}\rho$ | -1.62 | 3.33e-2 | 0.972 | normal | estimated |
| rate constant for conversion of target cells to a refractory state ($\log_{10}$ cell$^{-1}$day$^{-1}$ ) | $\log_{10}\phi$ | -5.29 | 6.52e-2 | 1.2 | normal | estimated |
| Deviation from $\log_{10}\phi$ for delta/other | $\beta_{\phi\_omicron}$ | 0.324 | 7.38e-2 | -- | -- | -- |
| Deviation from $\log_{10}\phi$ for delta/other | $\beta_{\phi\_none}$ | 1.53 | 0.134 | -- | -- | -- |
| infected cell clearance rate when I = 1 (day$^{-1}$ cells$^{-h}$) | $\delta$ | 1.38 | 2.64e-2 | 0.565 | log normal | estimated |
| onset of acquired immunity relative to detection (days) | $\tau$ | 15.9 | 0.453 | 0.461 | log normal | estimated |
| Deviation from $\tau$ for delta/other | $\beta_{\tau\_omicron}$ | -.364 | 3.53e-2 | -- | -- | -- |
| Deviation from $\tau$ for delta/other | $\beta_{\tau\_none}$ | -.422 | 6.14e-2 | -- | -- | -- |
| Deviation from $\tau$ for unvaccinated/no record | $\beta_{\tau\_>1\ vax}$ | -.141 | 3.16e-2 | -- | -- | -- |
| Increase in clearance rate of infected cells due to acquired immunity (day$^{-1}$) | $m$ | 16.4 | 0.542 | 0.502 | log normal | estimated |
| delay between infection in nasal tissue and detection (days) | $t_0$ | 2.08 | 2.8e-2 | 0.900 | logit [0,20] | estimated |
| viral clearance rate (day$^{-1}$) | $\gamma$ | 15 | -- | -- | -- | Goyal et al. |
| mean eclipse phase duration (days$^{-1}$) | $1/k$ | 1/4 | -- | -- | -- | Ke et al. |

Agreement between data and model-based quantitative features.

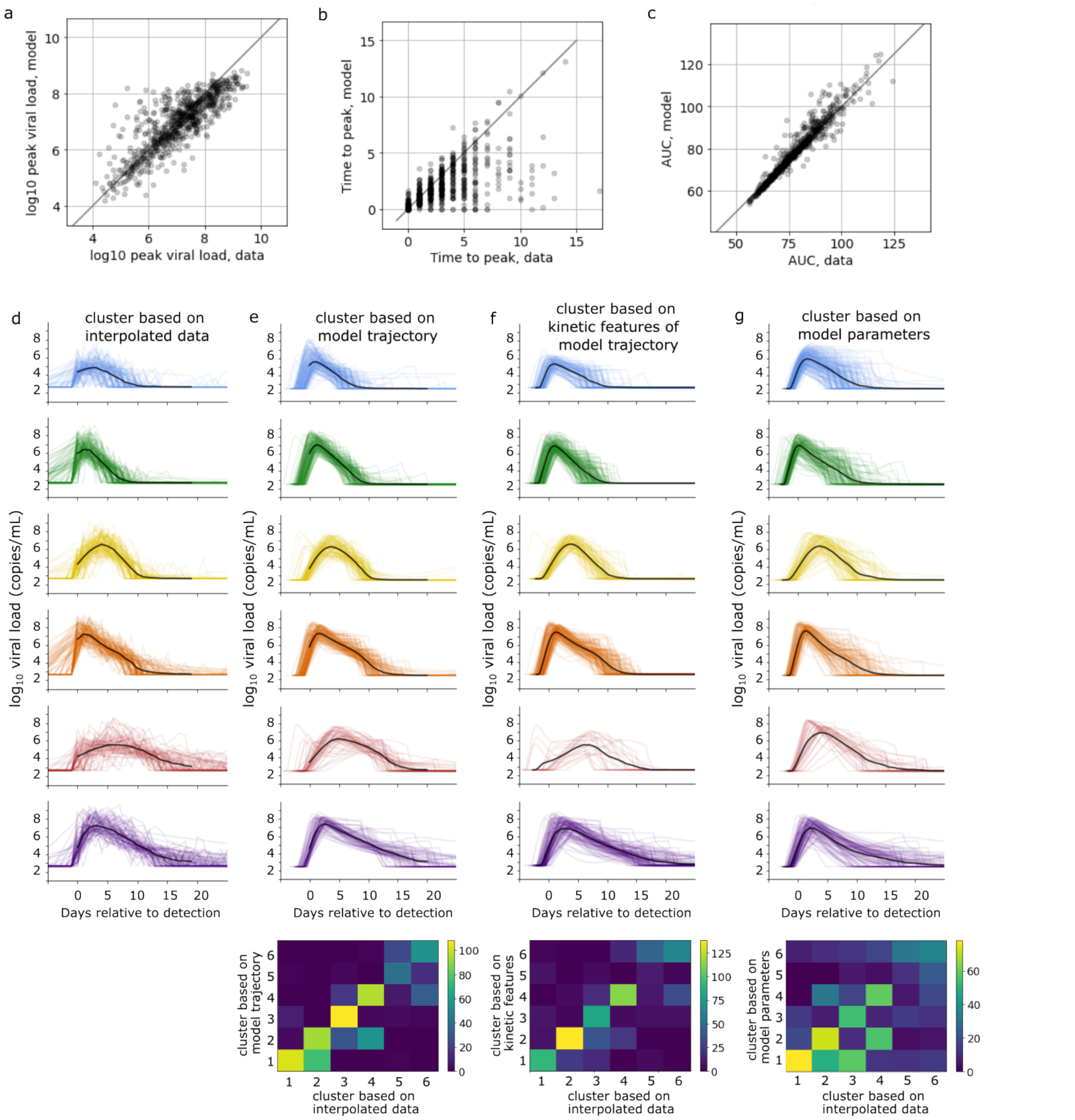


***Figure S1: Model validation.*** *We compared 3 three quantities computed from the data with estimates from model simulation. (a) There is reasonable agreement between peak viral loads. (b) The time to peak estimated by the model is often shorter than what is estimated from the data alone, as many true infections lack early measurements. (c) AUC predicted by the model and AUC predicted by the data have strong agreement (when we use the linearly interpolated data and sample modeled viral load daily). We compared results using k-means clustering with k =6 based on (d) time-series data from the NBA cohort against three other possible feature sets from model output: (e) viral load trajectories predicted by the model simulations: most cluster centers closely match their counterparts in (d), though group 1 had slightly earlier peak viral loads; the results agreed with the time-series data based clustering for 72% of cases. (f) viral kinetic features (peak, time to peak, duration, AUC) extracted from each simulation: again, a similar set of 6 characteristic clusters emerged; however, group 5 was smaller; clustering results agreed with the data-based clusters (d) in only 54% of cases. (g) normalized estimated model parameters as features; in this case, clusters did not agree as well with the clustering of the cohort data in (d). The bottom row shows degree of agreement with each column and the clustered data in (d).*

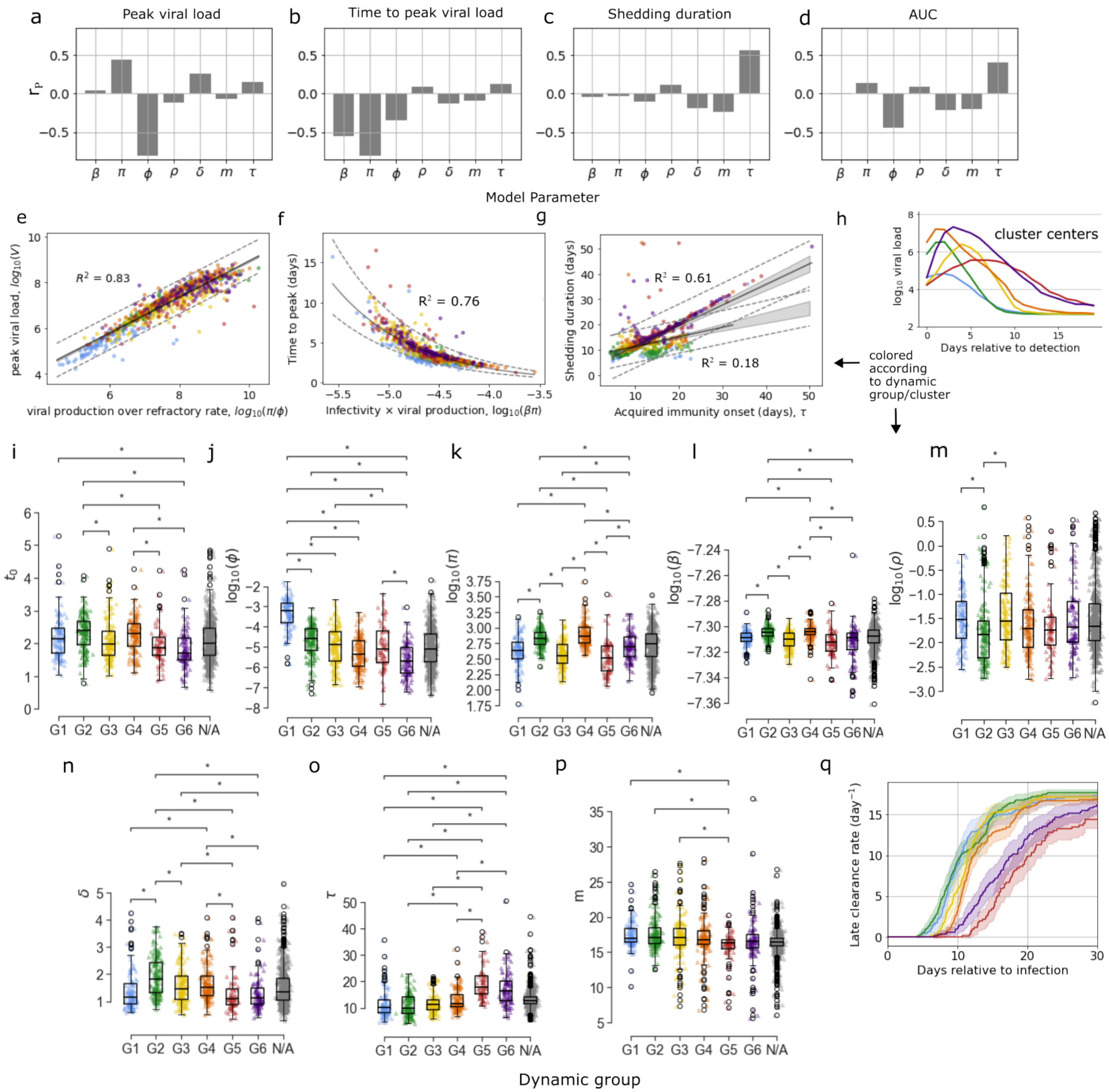


***Figure S2: Differences between dynamic clusters.*** *The first row shows the Pearson correlation coefficient, $r_p$, between model parameters and kinetic quantities (a) peak viral load, (b) time to peak viral load, (c) shedding duration and (d) area under the viral load curve. The second row shows how well combinations of parameters predict (e) peak viral load, (f) time to peak viral load, or (g) shedding duration. The coefficient of determination, $R^2$, is indicated, the best fit line is shown in black with a boot-strapped 95% confidence interval shaded, and the prediction interval is shown with dotted lines. For shedding duration, the plot shows predictors calculated separately with groups 1, 5 and 6 yielding $R^2 = 0.61$ and groups 2-4 yielding $R^2 = 0.18$. Points are colored according to their dynamic group, the centers of which are reproduced in panel (h). Panels (i-p) show the distributions of estimated individual parameter values stratified by dynamic group. Here pairs of groups indicated by a star have significantly different mean parameter values as identified by the Mann-Whitney U-test with $p_{adjusted} < .05$, where $p_{adjusted}$ is scaled using the Bonferroni correction for multiple comparisons. Panel (q) shows the average magnitude of the infected cell clearance rate due to delayed acquired immunity over time, stratified by dynamic group.*

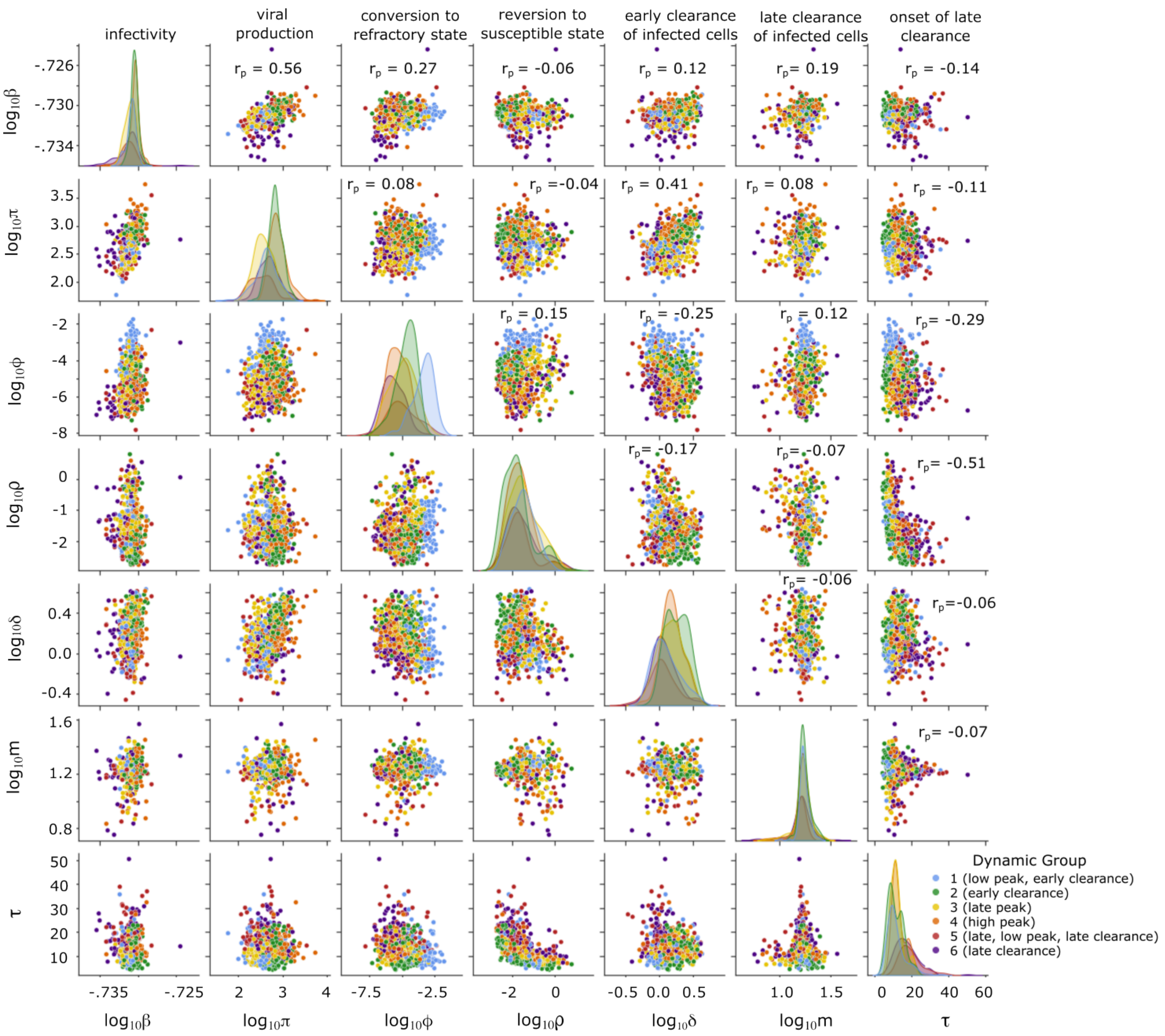


***Figure S3: Distributions of model parameters and pair-wise correlations.*** *Along the diagonal, the distribution of each parameter is shown for the six dynamic groups. Off-diagonal, for each pair of parameters, estimated individual parameter values are plotted with dots colored according to dynamic group. Panels are annotated with the Pearson correlation coefficient for that pair of parameters. The delay between infection and detection were excluded as it did not show strong correlations with other parameters.*

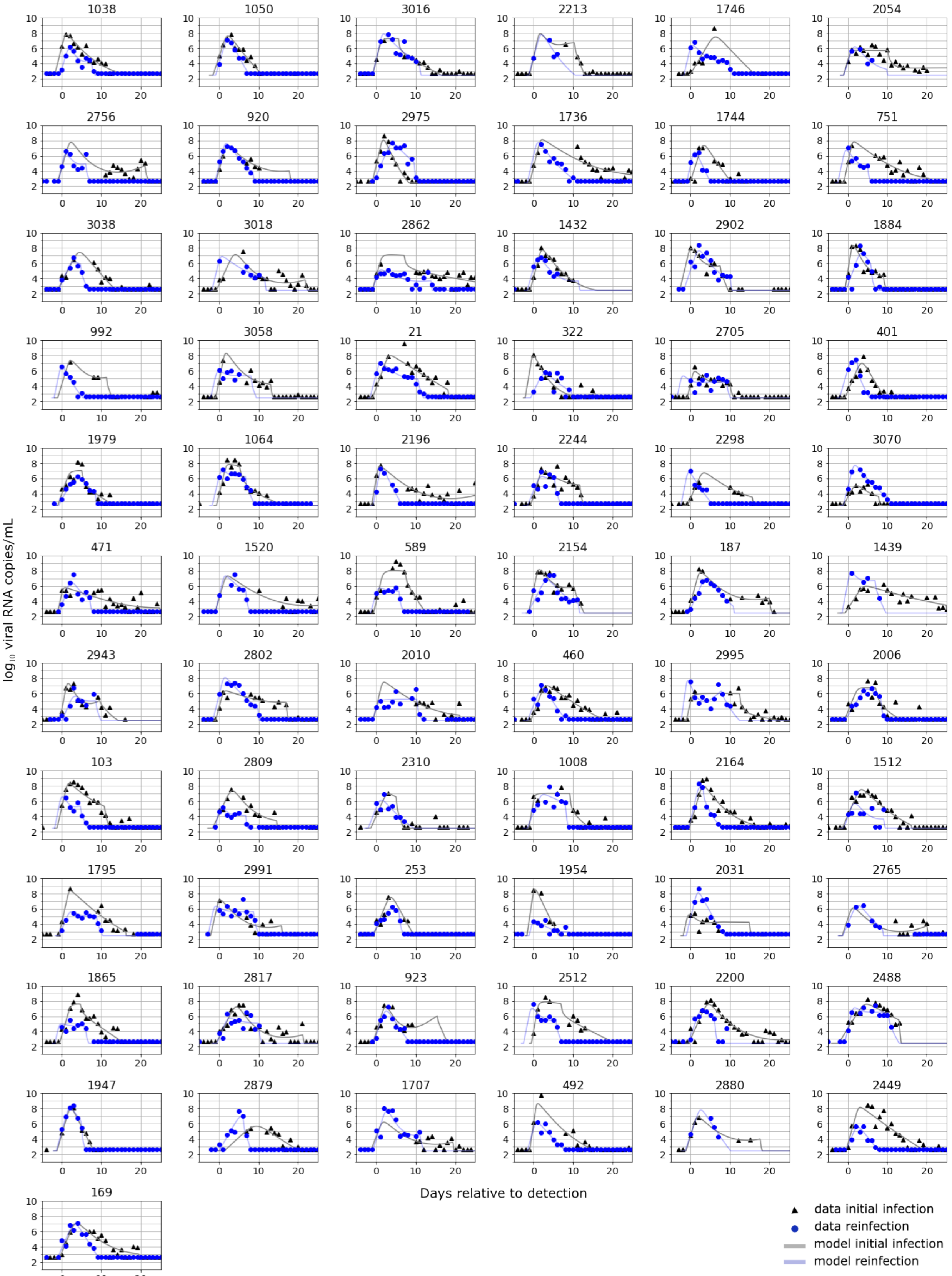


*Figure S4: 67 cases of primary infection and reinfection in the same individual documented in the NBA cohort.*

Tuning k-means clustering hyperparameters.

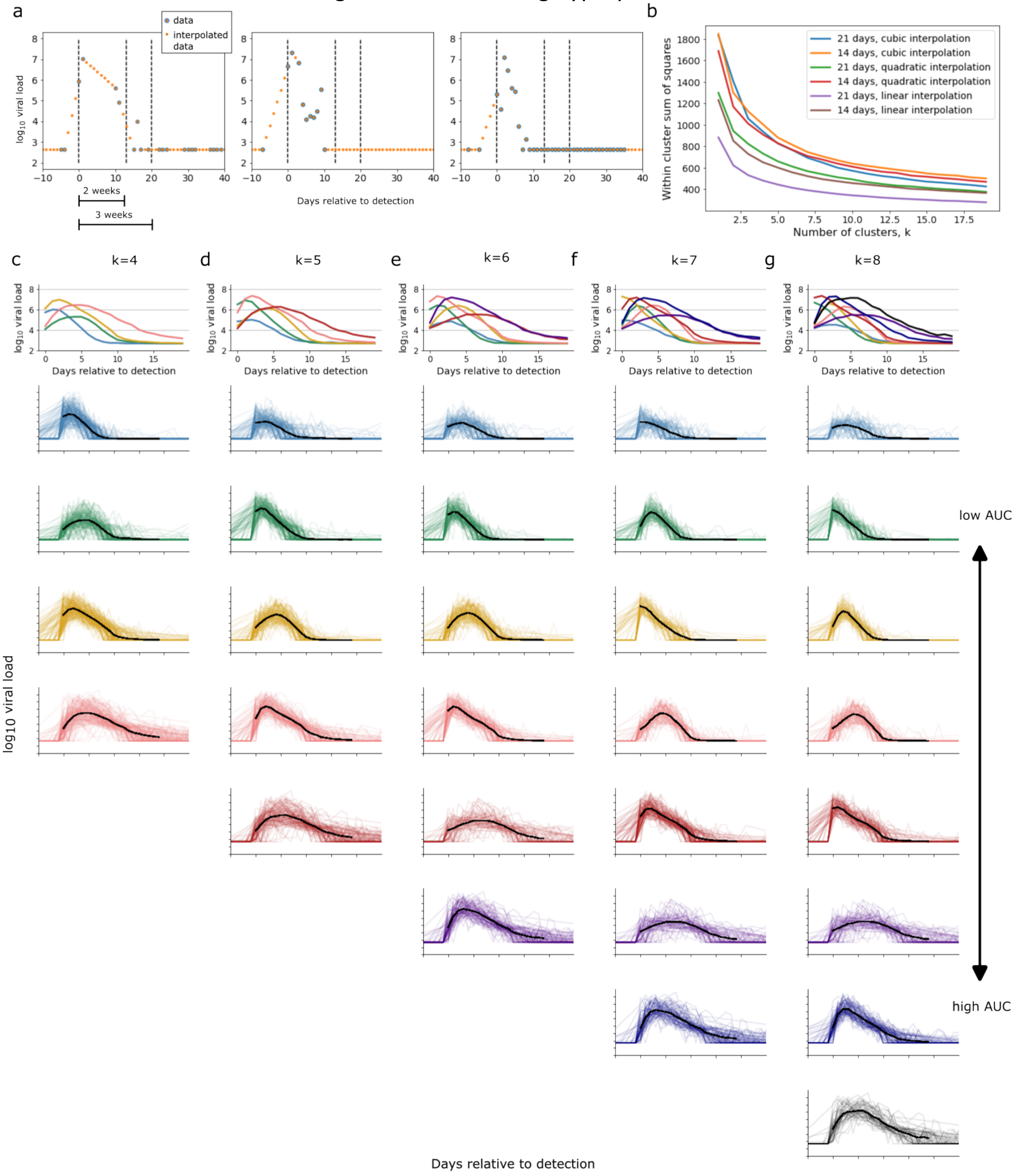


***Figure S5: Examining various cluster sizes.*** *We partitioned 768 infections from the NBA cohort that were documented over at least 3 weeks into subgroups with similar dynamics using k-means clustering. Before settling on using a 3-week documentation window and linear interpolation to impute any missing data points, we also considered a 2-week documentation window and interpolation using a quadratic or cubic spline. We examined the clustering results for k = 4 through k = 8, as this spanned the apparent "elbow" in the within-cluster sum of squares. The resulting clusters were ordered from low to high AUC for ease of comparison across scenarios.*

***Figure S6: Viral load data and predicted viral load for 1510 SARS-CoV-2 infections documented in the NBA cohort.*** *Cases of rebound are indicated with an R in the panel title and are included first. The number prior to the underscore corresponds to the person ID assigned by Hay et al. (eLife 2022) while the number after the underscore indicates whether this is a first, second, 3rd infection etc. The color indicates which dynamic group this data was assigned to by k-means clustering, as color-coded in Figures 2. Infections plotted in black/gray were excluded from clustering.*

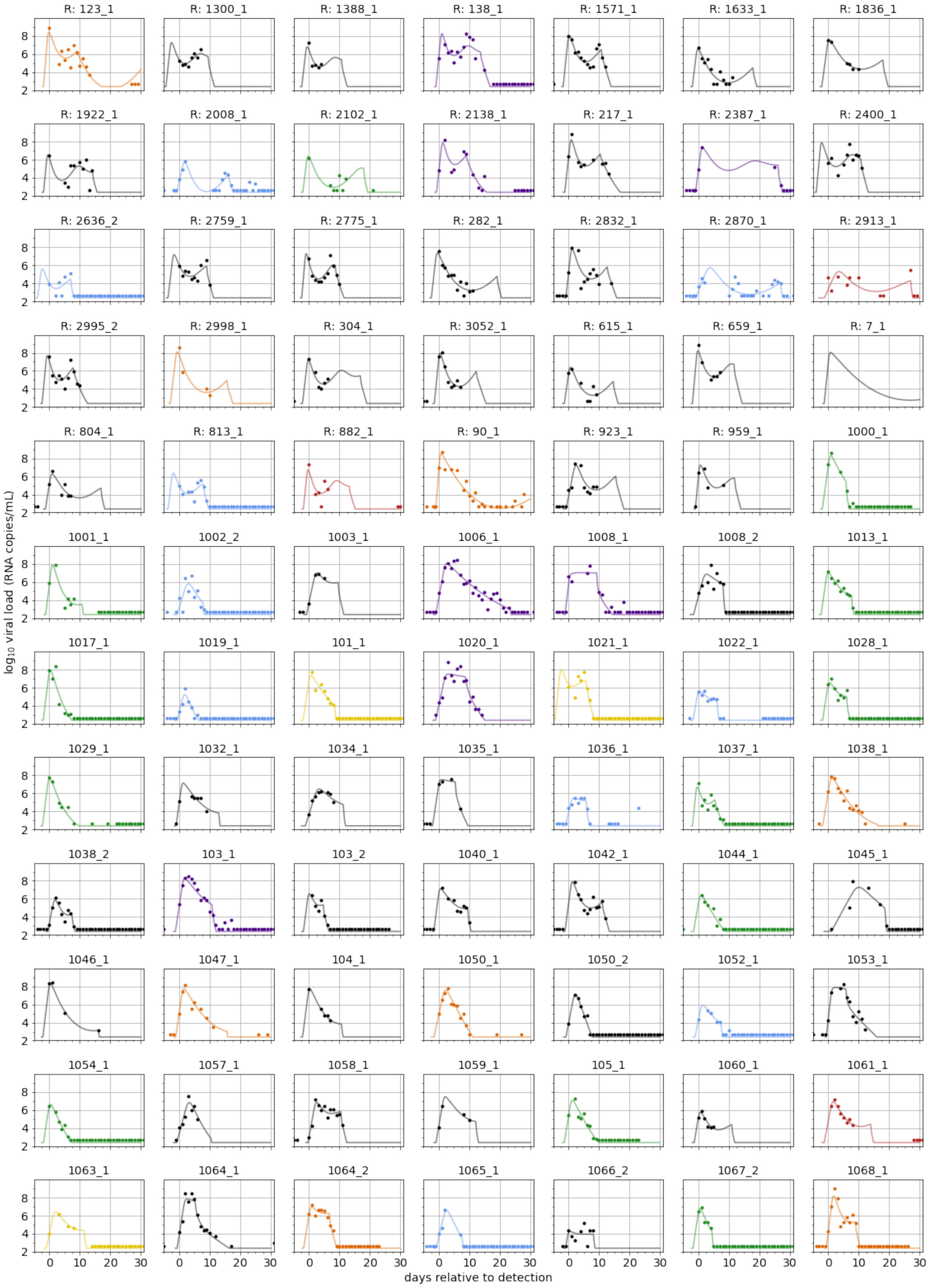
R: 123_1
R: 1300_1
R: 1388_1
R: 138_1
R: 1571_1
R: 1633_1
R: 1836_1
R: 1922_1
R: 2008_1
R: 2102_1
R: 2138_1
R: 217_1
R: 2387_1
R: 2400_1
R: 2636_2
R: 2759_1
R: 2775_1
R: 282_1
R: 2832_1
R: 2870_1
R: 2913_1
R: 2995_2
R: 2998_1
R: 304_1
R: 3052_1
R: 615_1
R: 659_1
R: 7_1
R: 804_1
R: 813_1
R: 882_1
R: 90_1
R: 923_1
R: 959_1
1000_1
1001_1
1002_2
1003_1
1006_1
1008_1
1008_2
1013_1
1017_1
1019_1
101_1
1020_1
1021_1
1022_1
1028_1
1029_1
1032_1
1034_1
1035_1
1036_1
1037_1
1038_1
1038_2
103_1
103_2
1040_1
1042_1
1044_1
1045_1
1046_1
1047_1
104_1
1050_1
1050_2
1052_1
1053_1
1054_1
1057_1
1058_1
1059_1
105_1
1060_1
1061_1
1063_1
1064_1
1064_2
1065_1
1066_2
1067_2
1068_1
log10 viral load (RNA copies/mL)
days relative to detection

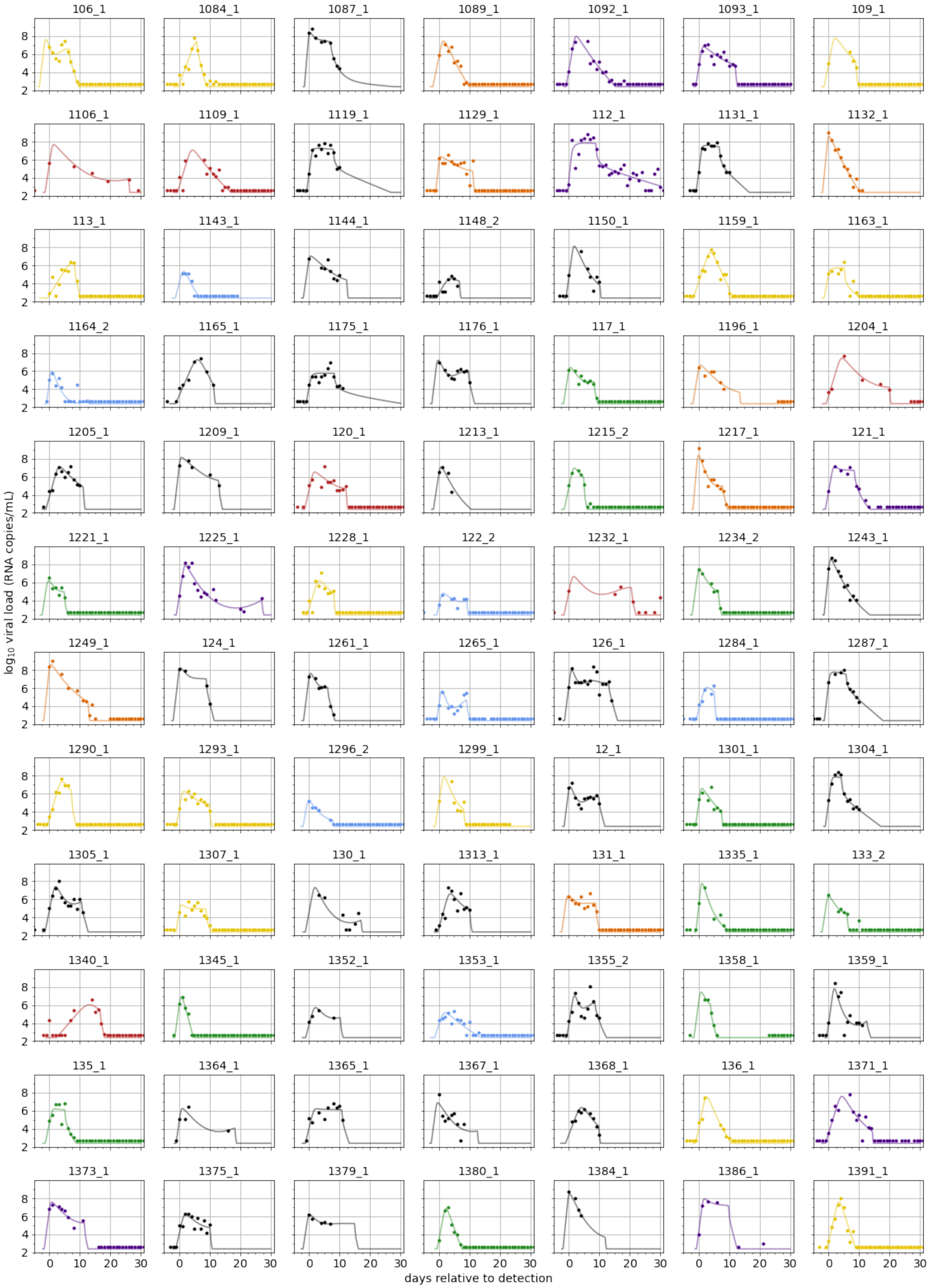
106_1
1084_1
1087_1
1089_1
1092_1
1093_1
109_1
1106_1
1109_1
1119_1
1129_1
112_1
1131_1
1132_1
113_1
1143_1
1144_1
1148_2
1150_1
1159_1
1163_1
1164_2
1165_1
1175_1
1176_1
117_1
1196_1
1204_1
1205_1
1209_1
120_1
1213_1
1215_2
1217_1
121_1
1221_1
1225_1
1228_1
122_2
1232_1
1234_2
1243_1
1249_1
124_1
1261_1
1265_1
126_1
1284_1
1287_1
1290_1
1293_1
1296_2
1299_1
12_1
1301_1
1304_1
1305_1
1307_1
130_1
1313_1
131_1
1335_1
133_2
1340_1
1345_1
1352_1
1353_1
1355_2
1358_1
1359_1
135_1
1364_1
1365_1
1367_1
1368_1
136_1
1371_1
1373_1
1375_1
1379_1
1380_1
1384_1
1386_1
1391_1
log10 viral load (RNA copies/mL)
days relative to detection

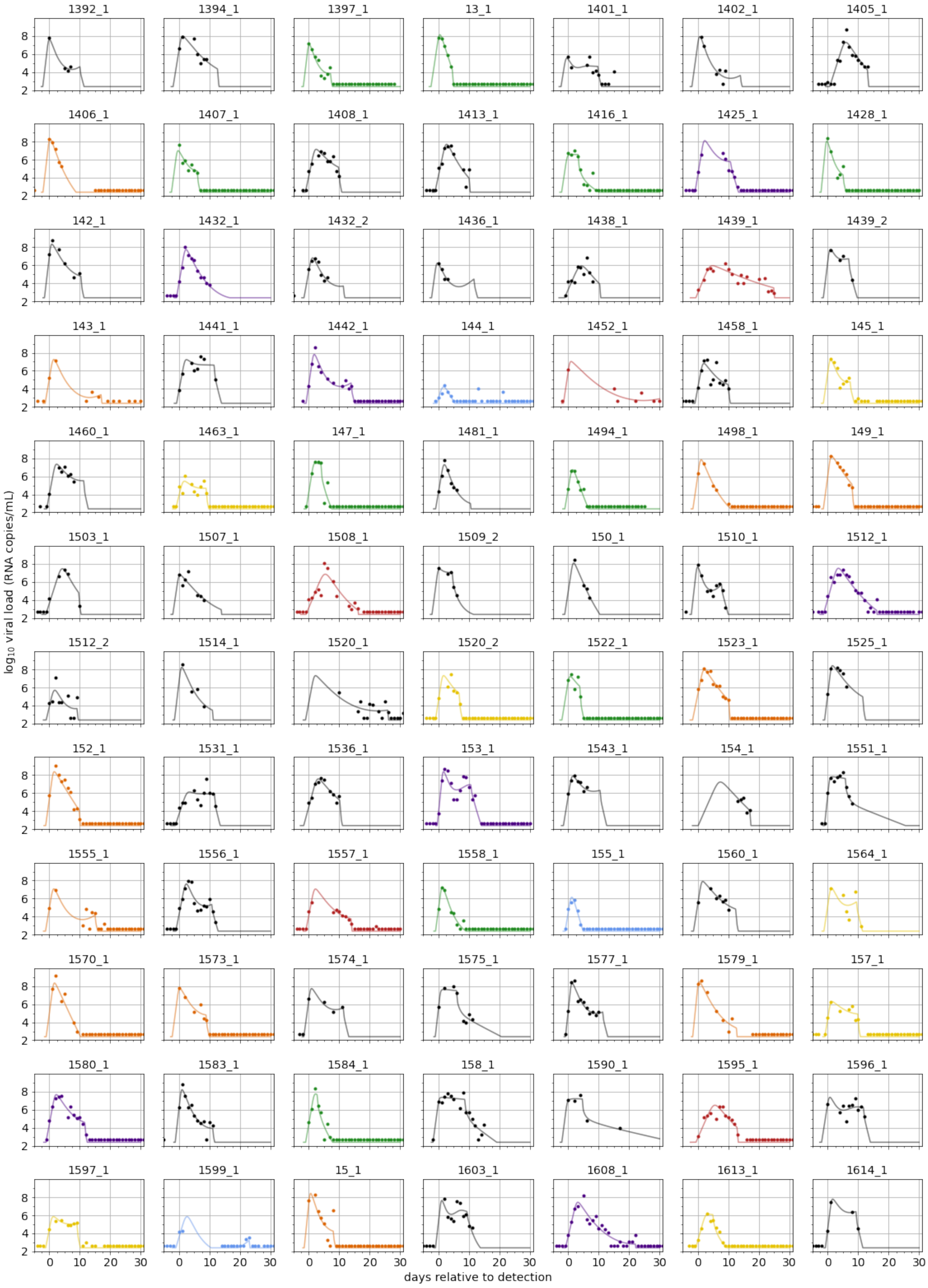
1392_1
1394_1
1397_1
13_1
1401_1
1402_1
1405_1
1406_1
1407_1
1408_1
1413_1
1416_1
1425_1
1428_1
142_1
1432_1
1432_2
1436_1
1438_1
1439_1
1439_2
143_1
1441_1
1442_1
144_1
1452_1
1458_1
145_1
1460_1
1463_1
147_1
1481_1
1494_1
1498_1
149_1
1503_1
1507_1
1508_1
1509_2
150_1
1510_1
1512_1
1512_2
1514_1
1520_1
1520_2
1522_1
1523_1
1525_1
152_1
1531_1
1536_1
153_1
1543_1
154_1
1551_1
1555_1
1556_1
1557_1
1558_1
155_1
1560_1
1564_1
1570_1
1573_1
1574_1
1575_1
1577_1
1579_1
157_1
1580_1
1583_1
1584_1
158_1
1590_1
1595_1
1596_1
1597_1
1599_1
15_1
1603_1
1608_1
1613_1
1614_1
log10 viral load (RNA copies/mL)
days relative to detection

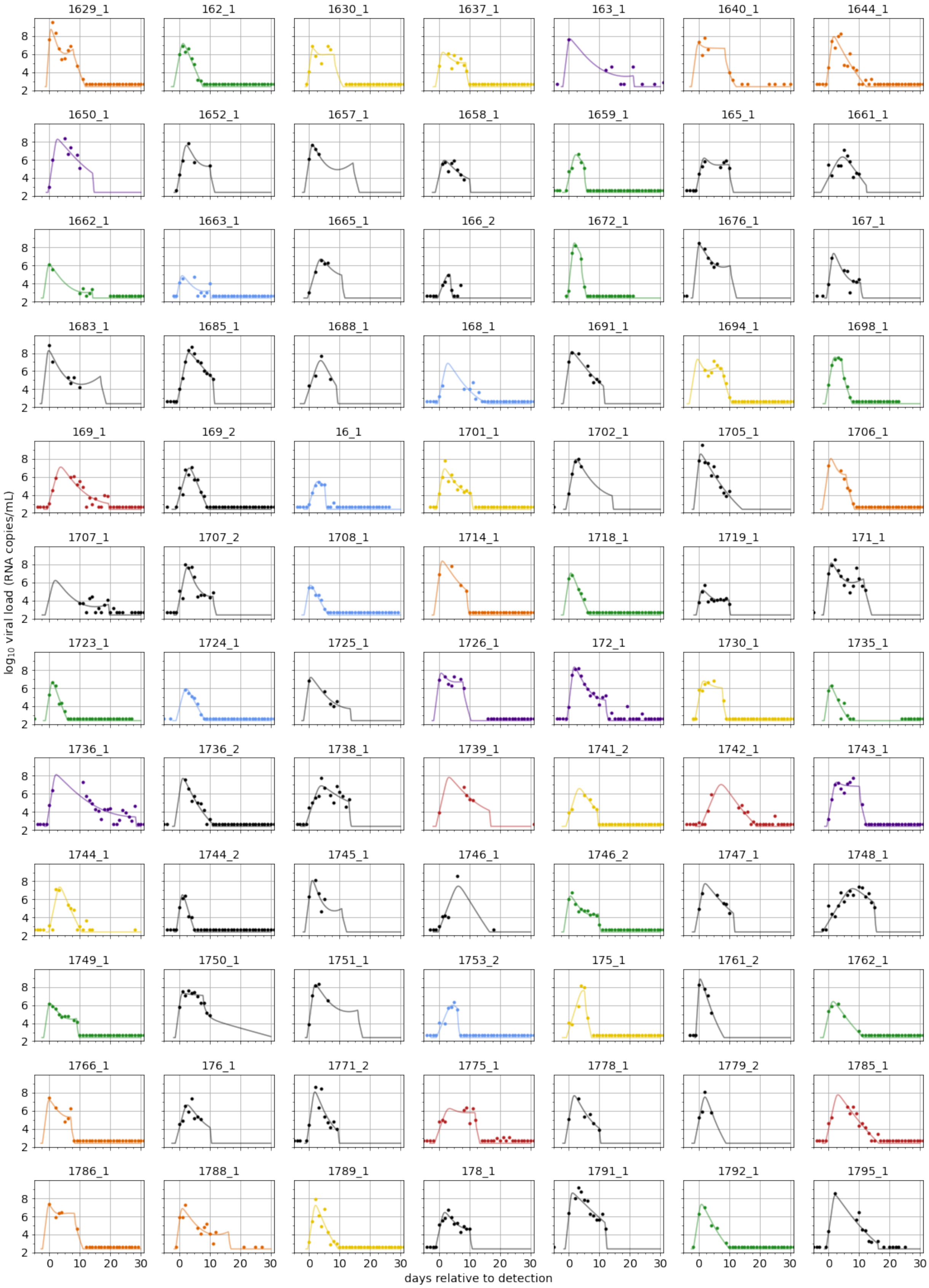
1629_1
162_1
1630_1
1637_1
163_1
1640_1
1644_1
1650_1
1652_1
1657_1
1658_1
1659_1
165_1
1661_1
1662_1
1663_1
1665_1
166_2
1672_1
1676_1
167_1
1683_1
1685_1
1688_1
168_1
1691_1
1694_1
1698_1
169_1
169_2
16_1
1701_1
1702_1
1705_1
1706_1
1707_1
1707_2
1708_1
1714_1
1718_1
1719_1
171_1
1723_1
1724_1
1725_1
1726_1
172_1
1730_1
1735_1
1736_1
1736_2
1738_1
1739_1
1741_2
1742_1
1743_1
1744_1
1744_2
1745_1
1746_1
1746_2
1747_1
1748_1
1749_1
1750_1
1751_1
1753_2
175_1
1761_2
1762_1
1766_1
176_1
1771_2
1775_1
1778_1
1779_2
1785_1
1786_1
1788_1
1789_1
178_1
1791_1
1792_1
1795_1
$\log_{10}$ viral load (RNA copies/mL)
days relative to detection

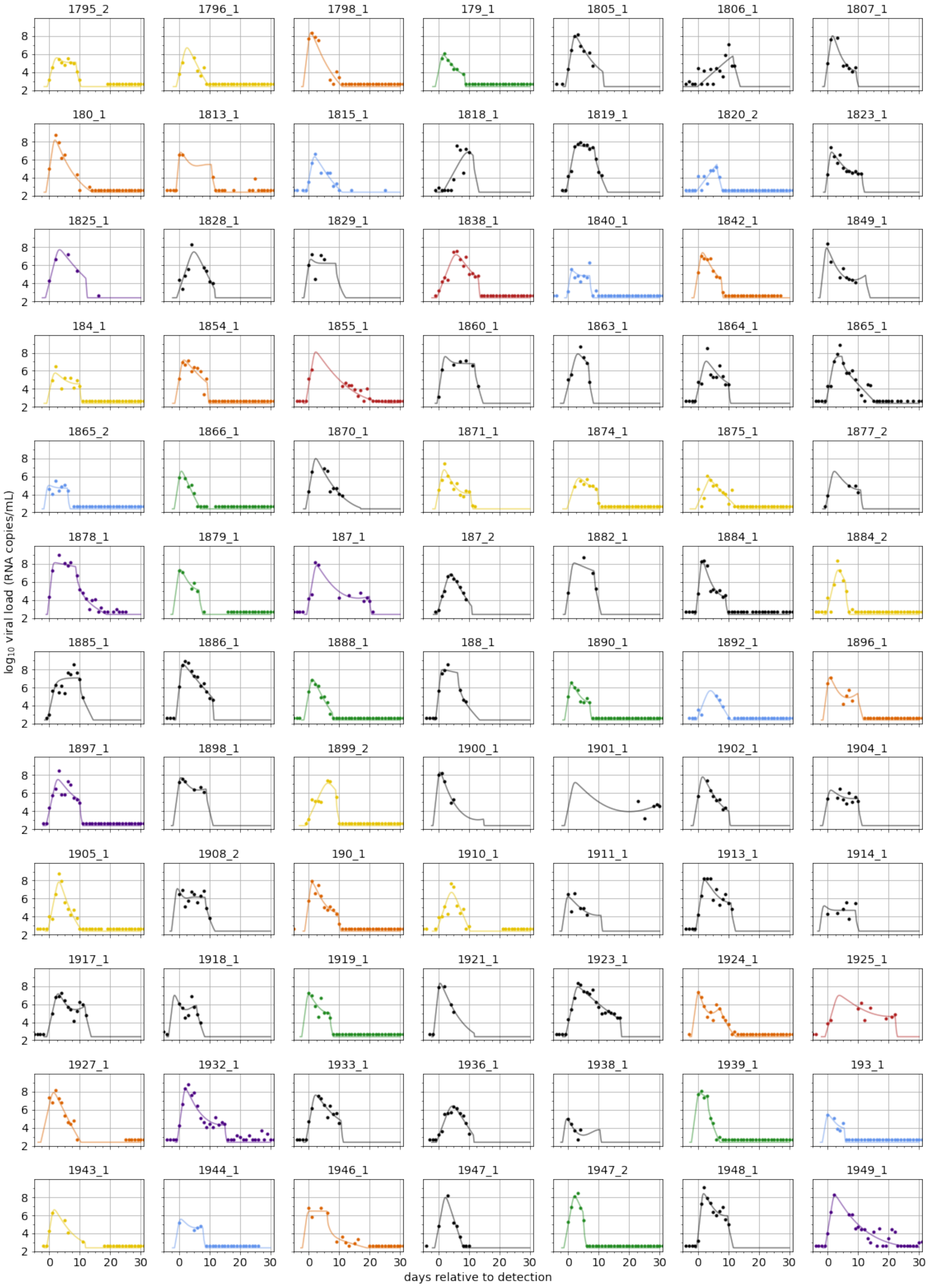
1795_2
1796_1
1798_1
179_1
1805_1
1806_1
1807_1
180_1
1813_1
1815_1
1818_1
1819_1
1820_2
1823_1
1825_1
1828_1
1829_1
1838_1
1840_1
1842_1
1849_1
184_1
1854_1
1855_1
1860_1
1863_1
1864_1
1865_1
1865_2
1866_1
1870_1
1871_1
1874_1
1875_1
1877_2
1878_1
1879_1
187_1
187_2
1882_1
1884_1
1884_2
1885_1
1886_1
1888_1
188_1
1890_1
1892_1
1896_1
1897_1
1898_1
1899_2
1900_1
1901_1
1902_1
1904_1
1905_1
1908_2
190_1
1910_1
1911_1
1913_1
1914_1
1917_1
1918_1
1919_1
1921_1
1923_1
1924_1
1925_1
1927_1
1932_1
1933_1
1936_1
1938_1
1939_1
193_1
1943_1
1944_1
1946_1
1947_1
1947_2
1948_1
1949_1
$\log_{10}$ viral load (RNA copies/mL)
days relative to detection

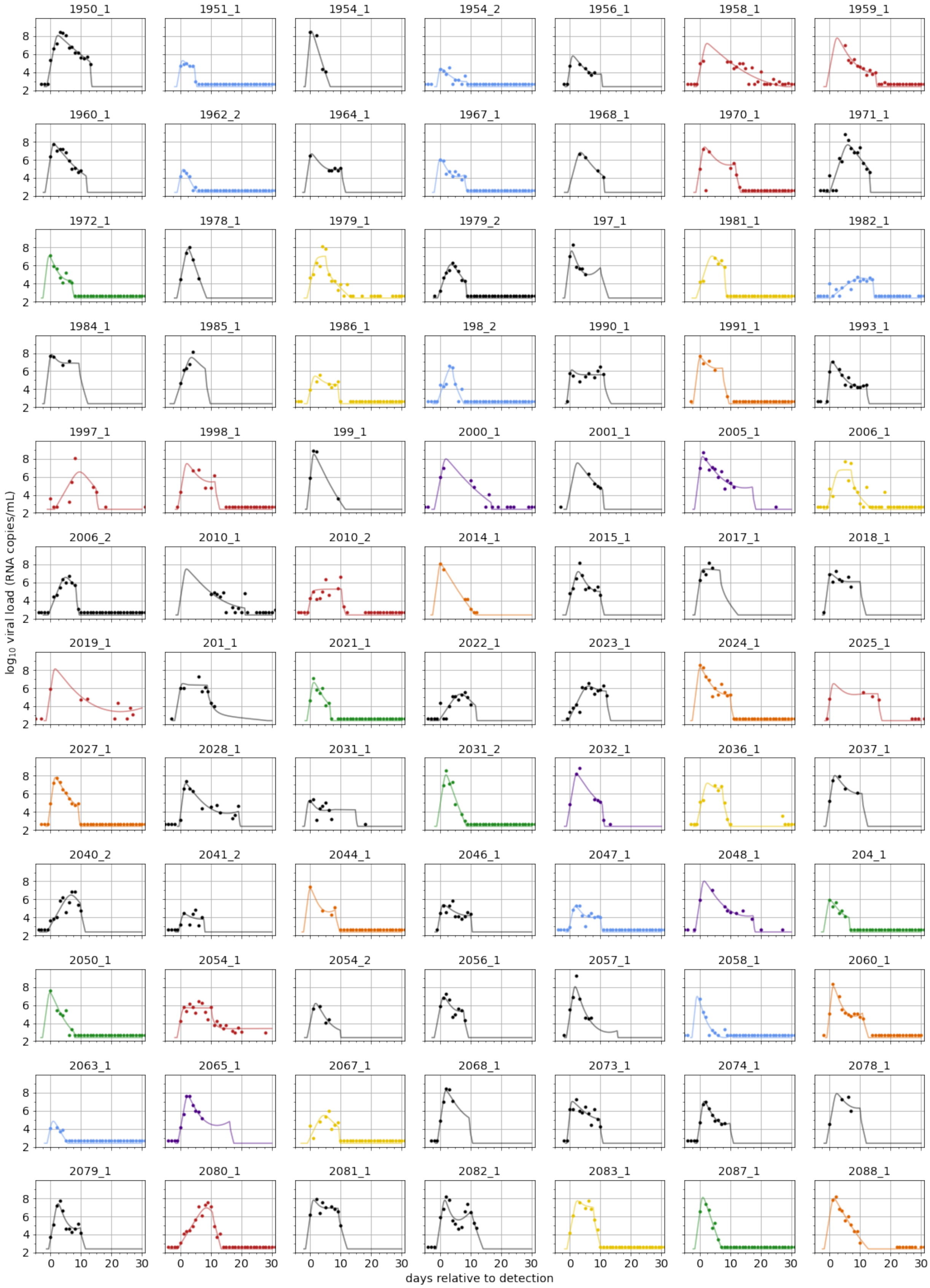
1950_1
1951_1
1954_1
1954_2
1956_1
1958_1
1959_1
1960_1
1962_2
1964_1
1967_1
1968_1
1970_1
1971_1
1972_1
1978_1
1979_1
1979_2
197_1
1981_1
1982_1
1984_1
1985_1
1986_1
198_2
1990_1
1991_1
1993_1
1997_1
1998_1
199_1
2000_1
2001_1
2005_1
2006_1
2006_2
2010_1
2010_2
2014_1
2015_1
2017_1
2018_1
2019_1
201_1
2021_1
2022_1
2023_1
2024_1
2025_1
2027_1
2028_1
2031_1
2031_2
2032_1
2036_1
2037_1
2040_2
2041_2
2044_1
2046_1
2047_1
2048_1
204_1
2050_1
2054_1
2054_2
2056_1
2057_1
2058_1
2060_1
2063_1
2065_1
2067_1
2068_1
2073_1
2074_1
2078_1
2079_1
2080_1
2081_1
2082_1
2083_1
2087_1
2088_1
log10 viral load (RNA copies/mL)
days relative to detection

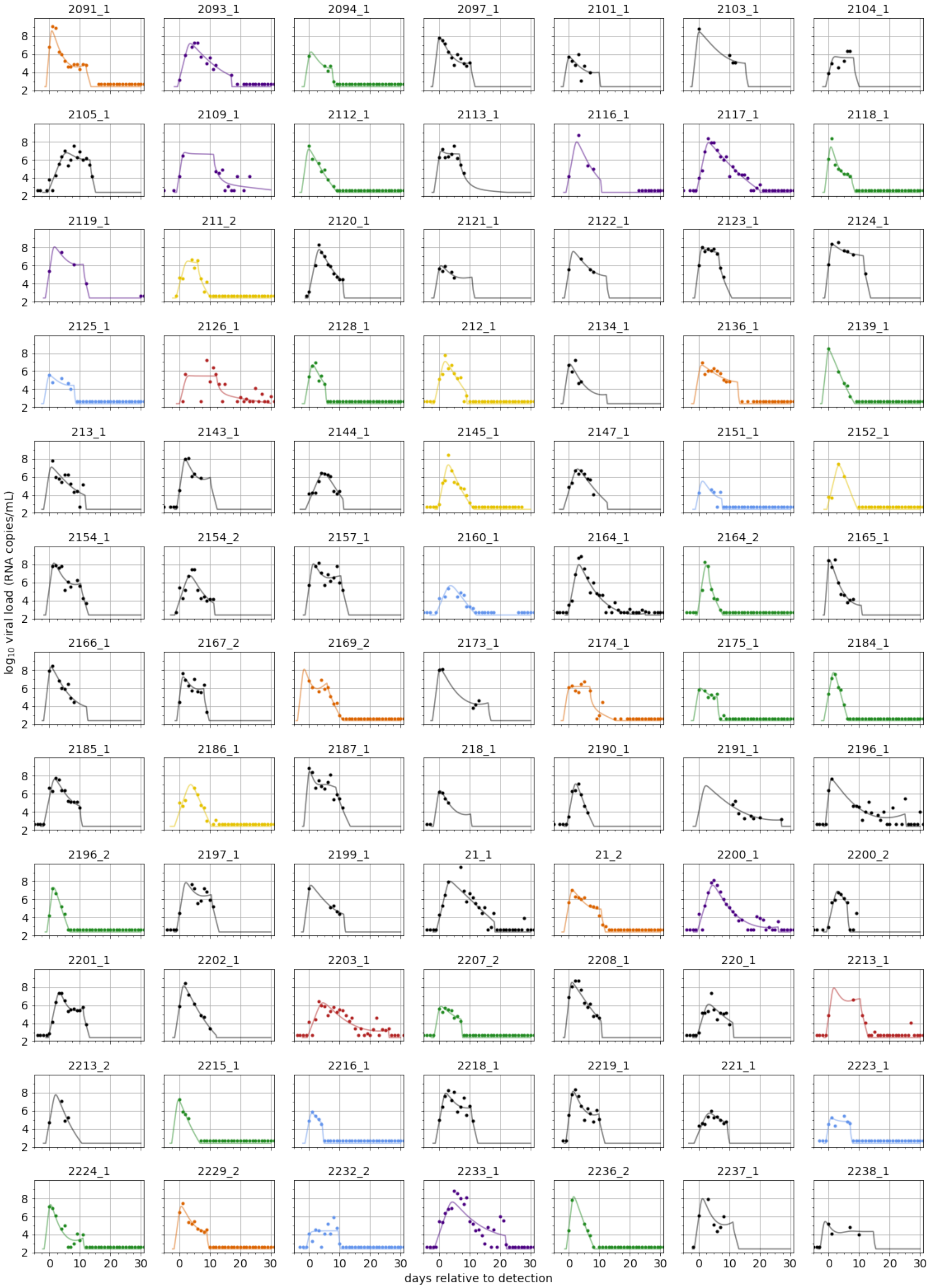
2091_1
2093_1
2094_1
2097_1
2101_1
2103_1
2104_1
2105_1
2109_1
2112_1
2113_1
2116_1
2117_1
2118_1
2119_1
211_2
2120_1
2121_1
2122_1
2123_1
2124_1
2125_1
2126_1
2128_1
212_1
2134_1
2136_1
2139_1
213_1
2143_1
2144_1
2145_1
2147_1
2151_1
2152_1
2154_1
2154_2
2157_1
2160_1
2164_1
2164_2
2165_1
2166_1
2167_2
2169_2
2173_1
2174_1
2175_1
2184_1
2185_1
2186_1
2187_1
218_1
2190_1
2191_1
2196_1
2196_2
2197_1
2199_1
21_1
21_2
2200_1
2200_2
2201_1
2202_1
2203_1
2207_2
2208_1
220_1
2213_1
2213_2
2215_1
2216_1
2218_1
2219_1
221_1
2223_1
2224_1
2229_2
2232_2
2233_1
2236_2
2237_1
2238_1
$\log_{10}$ viral load (RNA copies/mL)
days relative to detection

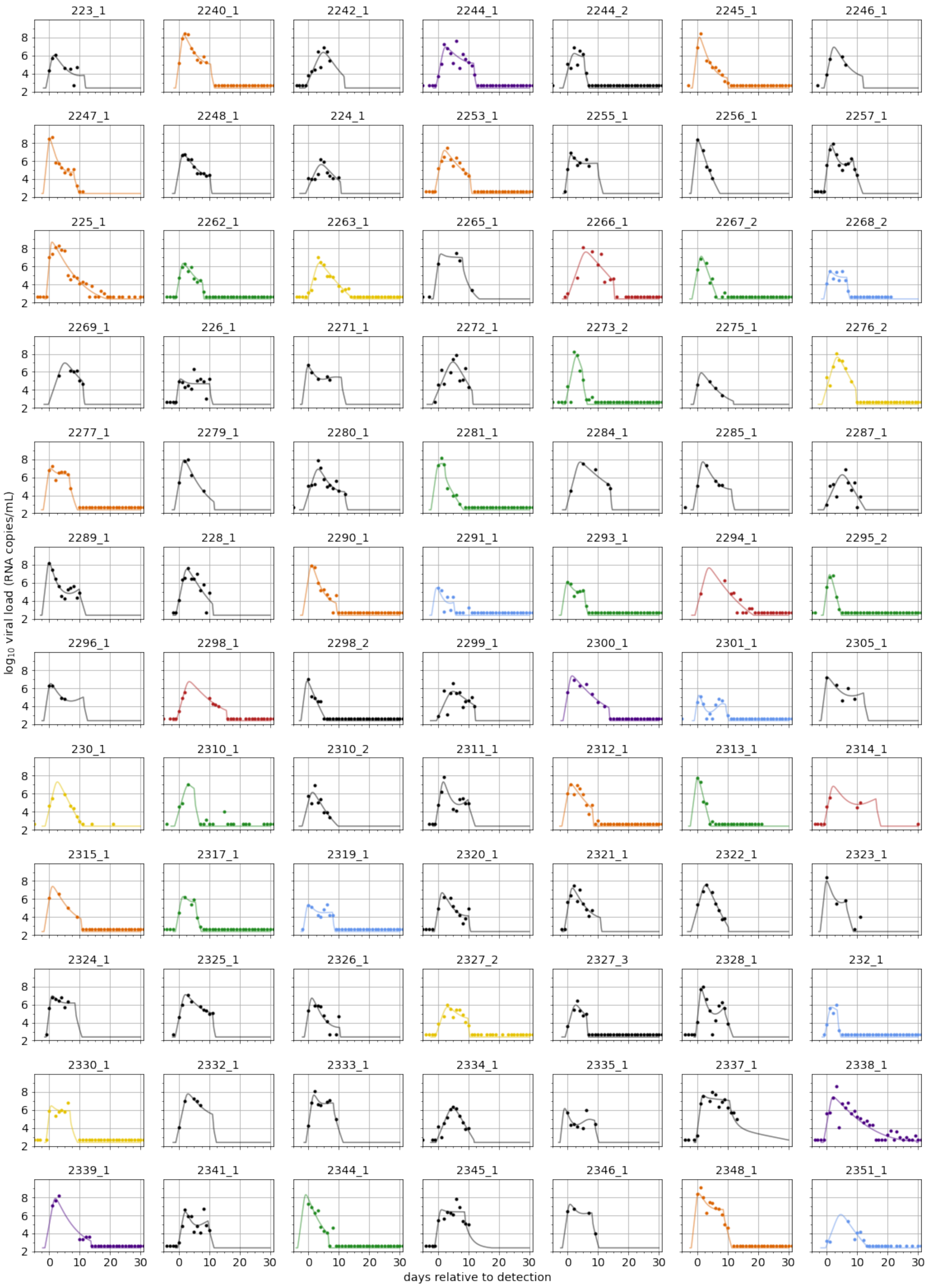
223_1
2240_1
2242_1
2244_1
2244_2
2245_1
2246_1
2247_1
2248_1
224_1
2253_1
2255_1
2256_1
2257_1
225_1
2262_1
2263_1
2265_1
2266_1
2267_2
2268_2
2269_1
226_1
2271_1
2272_1
2273_2
2275_1
2276_2
2277_1
2279_1
2280_1
2281_1
2284_1
2285_1
2287_1
2289_1
228_1
2290_1
2291_1
2293_1
2294_1
2295_2
2296_1
2298_1
2298_2
2299_1
2300_1
2301_1
2305_1
230_1
2310_1
2310_2
2311_1
2312_1
2313_1
2314_1
2315_1
2317_1
2319_1
2320_1
2321_1
2322_1
2323_1
2324_1
2325_1
2326_1
2327_2
2327_3
2328_1
232_1
2330_1
2332_1
2333_1
2334_1
2335_1
2337_1
2338_1
2339_1
2341_1
2344_1
2345_1
2346_1
2348_1
2351_1
$\log_{10}$ viral load (RNA copies/mL)
days relative to detection

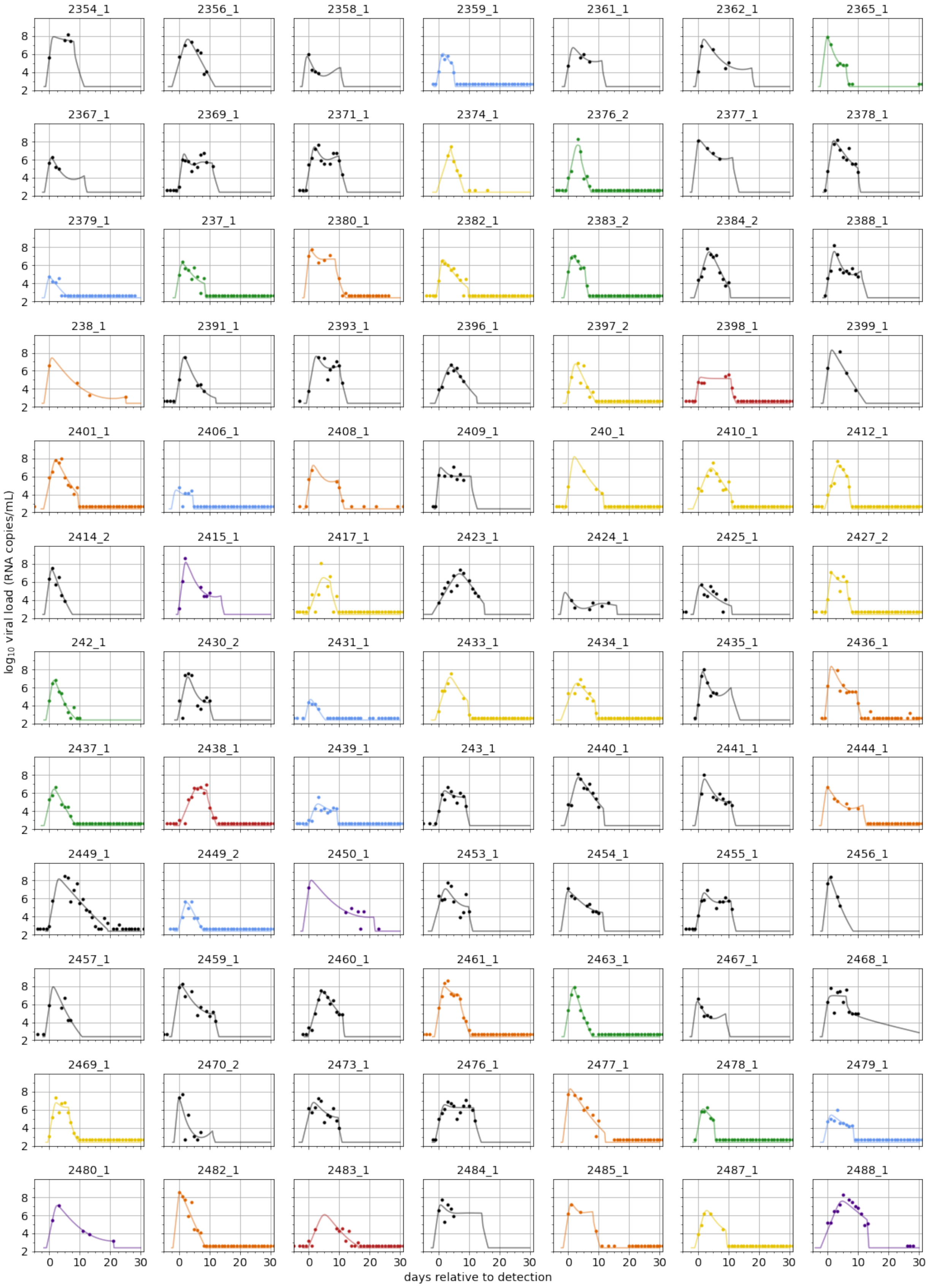
2354_1
2356_1
2358_1
2359_1
2361_1
2362_1
2365_1
2367_1
2369_1
2371_1
2374_1
2376_2
2377_1
2378_1
2379_1
237_1
2380_1
2382_1
2383_2
2384_2
2388_1
238_1
2391_1
2393_1
2396_1
2397_2
2398_1
2399_1
2401_1
2406_1
2408_1
2409_1
240_1
2410_1
2412_1
2414_2
2415_1
2417_1
2423_1
2424_1
2425_1
2427_2
242_1
2430_2
2431_1
2433_1
2434_1
2435_1
2436_1
2437_1
2438_1
2439_1
243_1
2440_1
2441_1
2444_1
2449_1
2449_2
2450_1
2453_1
2454_1
2455_1
2456_1
2457_1
2459_1
2460_1
2461_1
2463_1
2467_1
2468_1
2469_1
2470_2
2473_1
2476_1
2477_1
2478_1
2479_1
2480_1
2482_1
2483_1
2484_1
2485_1
2487_1
2488_1
$\log_{10}$ viral load (RNA copies/mL)
days relative to detection

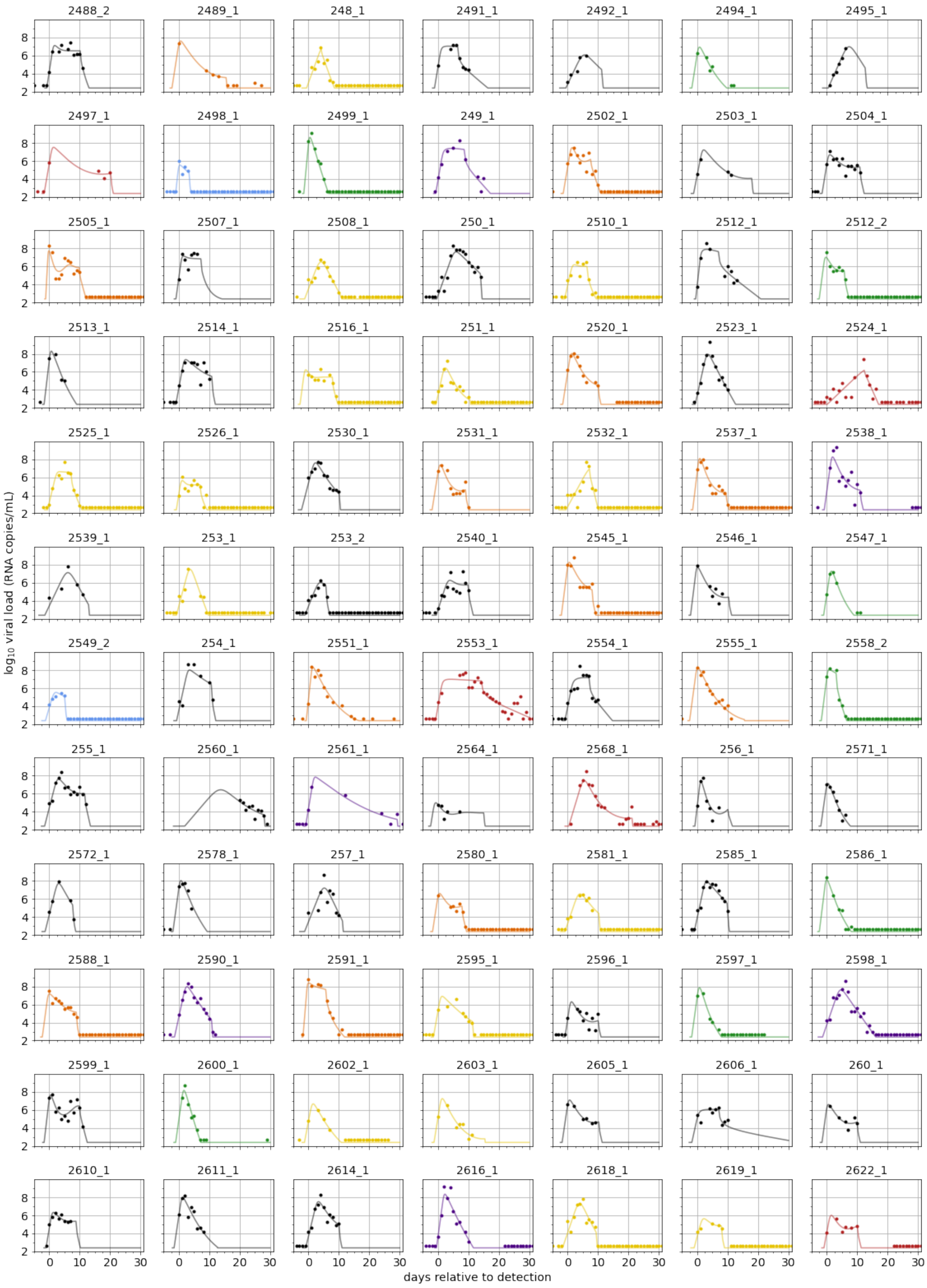
2488_2
2489_1
248_1
2491_1
2492_1
2494_1
2495_1
2497_1
2498_1
2499_1
249_1
2502_1
2503_1
2504_1
2505_1
2507_1
2508_1
250_1
2510_1
2512_1
2512_2
2513_1
2514_1
2516_1
251_1
2520_1
2523_1
2524_1
2525_1
2526_1
2530_1
2531_1
2532_1
2537_1
2538_1
2539_1
253_1
253_2
2540_1
2545_1
2546_1
2547_1
2549_2
254_1
2551_1
2553_1
2554_1
2555_1
2558_2
255_1
2560_1
2561_1
2564_1
2568_1
256_1
2571_1
2572_1
2578_1
257_1
2580_1
2581_1
2585_1
2586_1
2588_1
2590_1
2591_1
2595_1
2596_1
2597_1
2598_1
2599_1
2600_1
2602_1
2603_1
2605_1
2606_1
260_1
2610_1
2611_1
2614_1
2616_1
2618_1
2619_1
2622_1
log10 viral load (RNA copies/mL)
days relative to detection

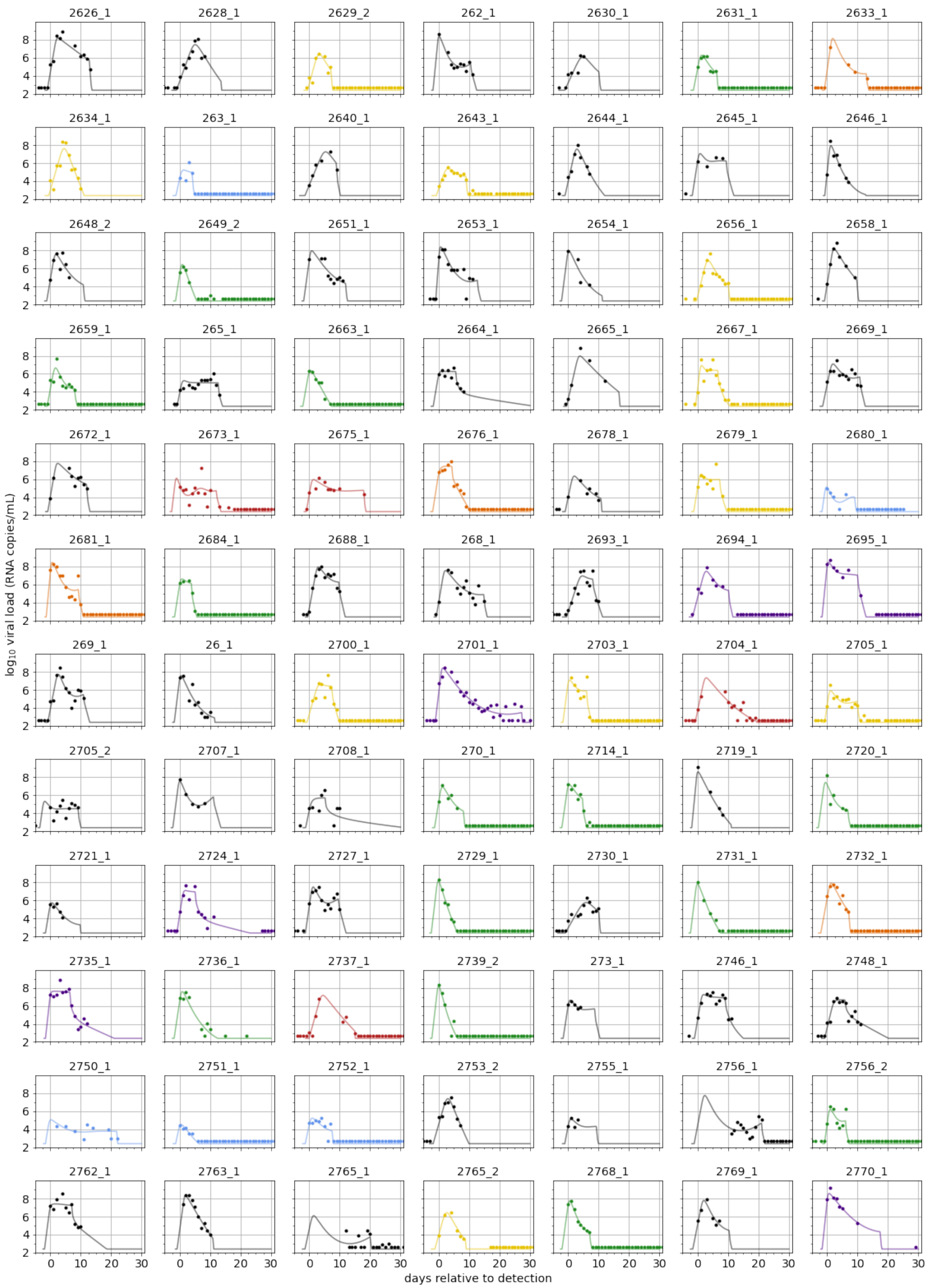
2626_1
2628_1
2629_2
262_1
2630_1
2631_1
2633_1
2634_1
263_1
2640_1
2643_1
2644_1
2645_1
2646_1
2648_2
2649_2
2651_1
2653_1
2654_1
2656_1
2658_1
2659_1
265_1
2663_1
2664_1
2665_1
2667_1
2669_1
2672_1
2673_1
2675_1
2676_1
2678_1
2679_1
2680_1
2681_1
2684_1
2688_1
268_1
2693_1
2694_1
2695_1
269_1
26_1
2700_1
2701_1
2703_1
2704_1
2705_1
2705_2
2707_1
2708_1
270_1
2714_1
2719_1
2720_1
2721_1
2724_1
2727_1
2729_1
2730_1
2731_1
2732_1
2735_1
2736_1
2737_1
2739_2
273_1
2746_1
2748_1
2750_1
2751_1
2752_1
2753_2
2755_1
2756_1
2756_2
2762_1
2763_1
2765_1
2765_2
2768_1
2769_1
2770_1
$\log_{10}$ viral load (RNA copies/mL)
days relative to detection

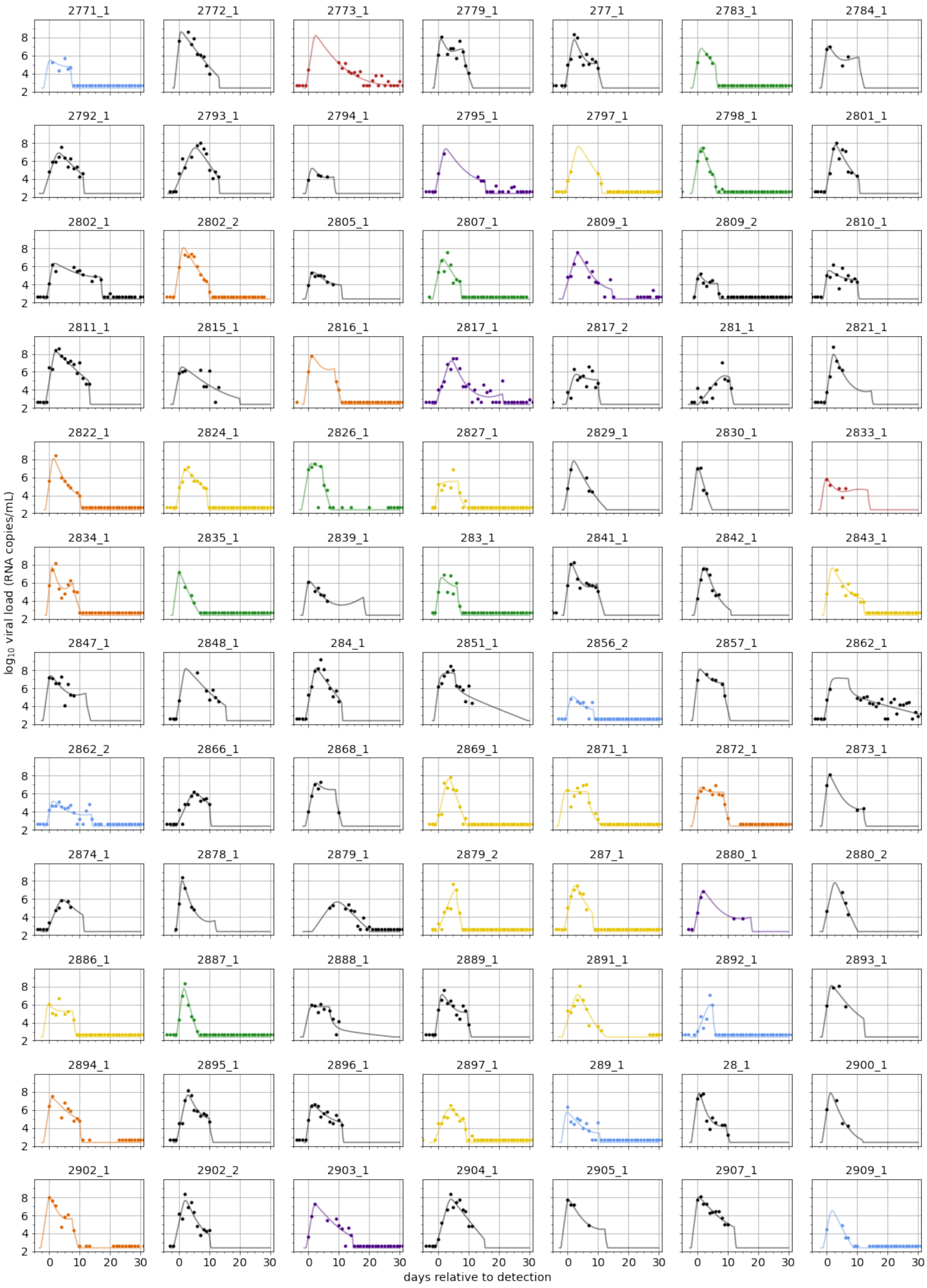
2771_1
2772_1
2773_1
2779_1
277_1
2783_1
2784_1
2792_1
2793_1
2794_1
2795_1
2797_1
2798_1
2801_1
2802_1
2802_2
2805_1
2807_1
2809_1
2809_2
2810_1
2811_1
2815_1
2816_1
2817_1
2817_2
281_1
2821_1
2822_1
2824_1
2826_1
2827_1
2829_1
2830_1
2833_1
2834_1
2835_1
2839_1
283_1
2841_1
2842_1
2843_1
2847_1
2848_1
284_1
2851_1
2856_2
2857_1
2862_1
2862_2
2866_1
2868_1
2869_1
2871_1
2872_1
2873_1
2874_1
2878_1
2879_1
2879_2
287_1
2880_1
2880_2
2886_1
2887_1
2888_1
2889_1
2891_1
2892_1
2893_1
2894_1
2895_1
2896_1
2897_1
289_1
28_1
2900_1
2902_1
2902_2
2903_1
2904_1
2905_1
2907_1
2909_1
$\log_{10}$ viral load (RNA copies/mL)
days relative to detection

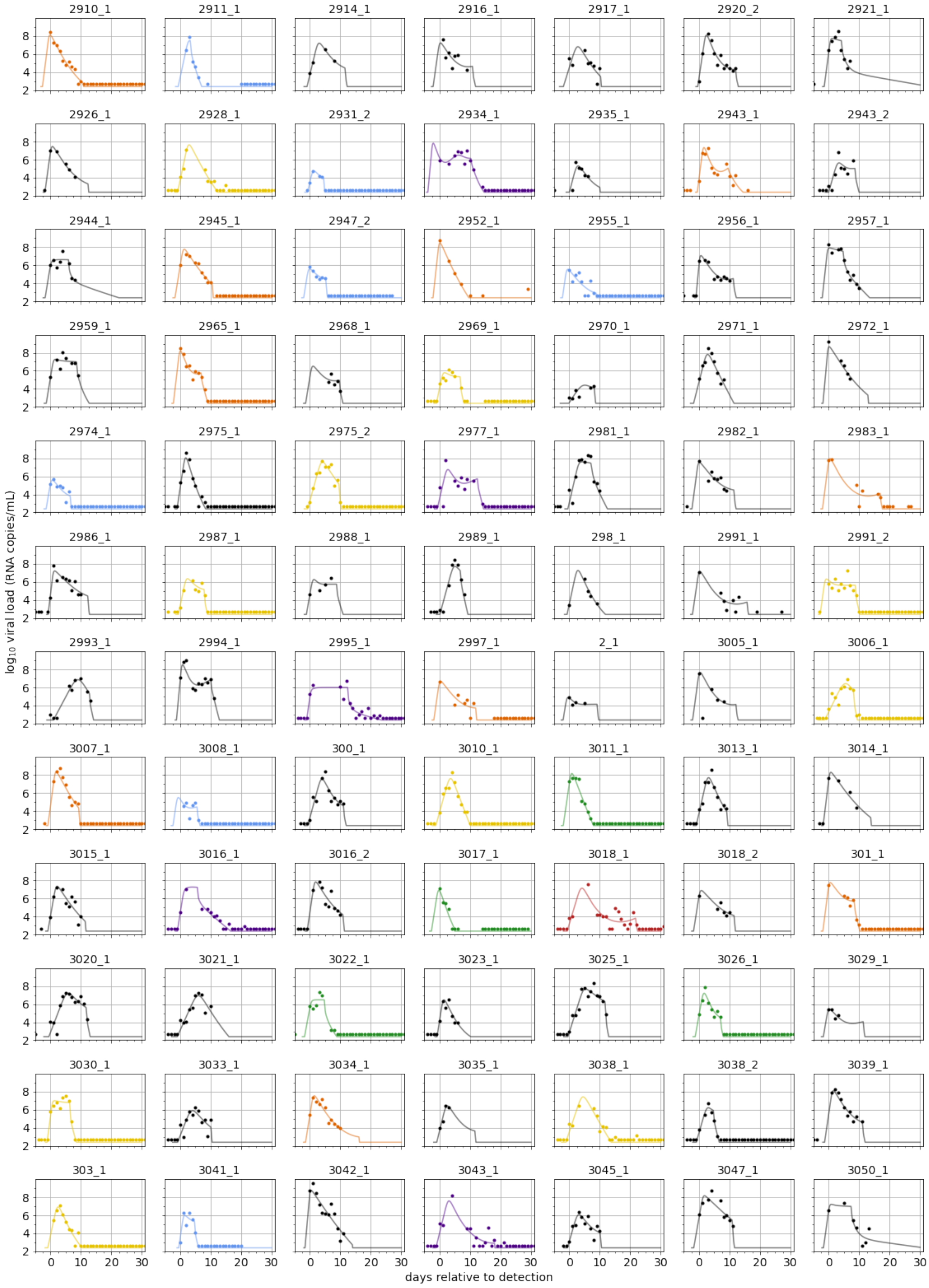
2910_1
2911_1
2914_1
2916_1
2917_1
2920_2
2921_1
2926_1
2928_1
2931_2
2934_1
2935_1
2943_1
2943_2
2944_1
2945_1
2947_2
2952_1
2955_1
2956_1
2957_1
2959_1
2965_1
2968_1
2969_1
2970_1
2971_1
2972_1
2974_1
2975_1
2975_2
2977_1
2981_1
2982_1
2983_1
2986_1
2987_1
2988_1
2989_1
298_1
2991_1
2991_2
2993_1
2994_1
2995_1
2997_1
2_1
3005_1
3006_1
3007_1
3008_1
300_1
3010_1
3011_1
3013_1
3014_1
3015_1
3016_1
3016_2
3017_1
3018_1
3018_2
301_1
3020_1
3021_1
3022_1
3023_1
3025_1
3026_1
3029_1
3030_1
3033_1
3034_1
3035_1
3038_1
3038_2
3039_1
303_1
3041_1
3042_1
3043_1
3045_1
3047_1
3050_1
$\log_{10}$ viral load (RNA copies/mL)
days relative to detection

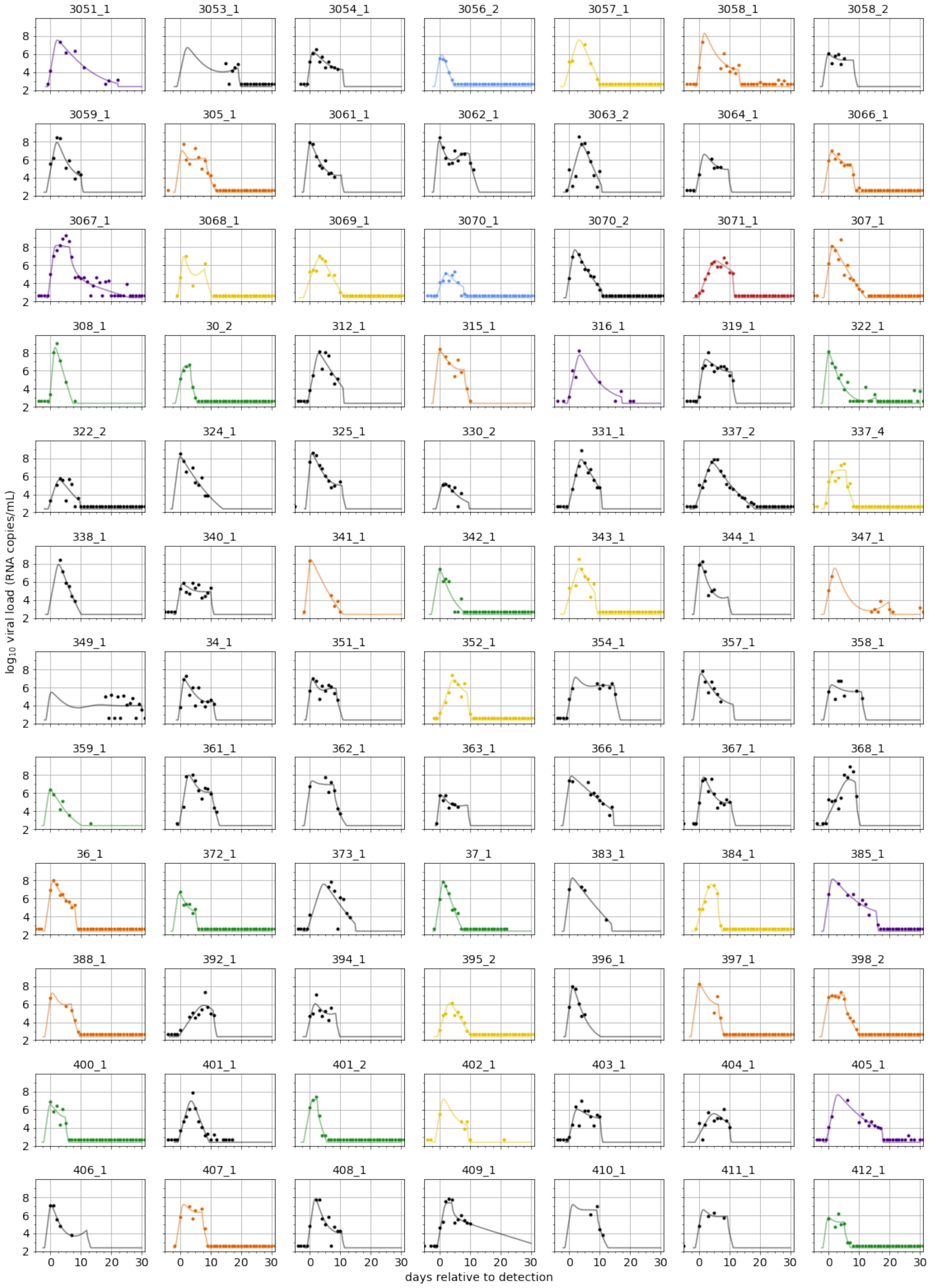
3051_1
3053_1
3054_1
3056_2
3057_1
3058_1
3058_2
3059_1
305_1
3061_1
3062_1
3063_2
3064_1
3066_1
3067_1
3068_1
3069_1
3070_1
3070_2
3071_1
307_1
308_1
30_2
312_1
315_1
316_1
319_1
322_1
322_2
324_1
325_1
330_2
331_1
337_2
337_4
338_1
340_1
341_1
342_1
343_1
344_1
347_1
349_1
34_1
351_1
352_1
354_1
357_1
358_1
359_1
361_1
362_1
363_1
366_1
367_1
368_1
36_1
372_1
373_1
37_1
383_1
384_1
385_1
388_1
392_1
394_1
395_2
396_1
397_1
398_2
400_1
401_1
401_2
402_1
403_1
404_1
405_1
406_1
407_1
408_1
409_1
410_1
411_1
412_1
$\log_{10}$ viral load (RNA copies/mL)
days relative to detection

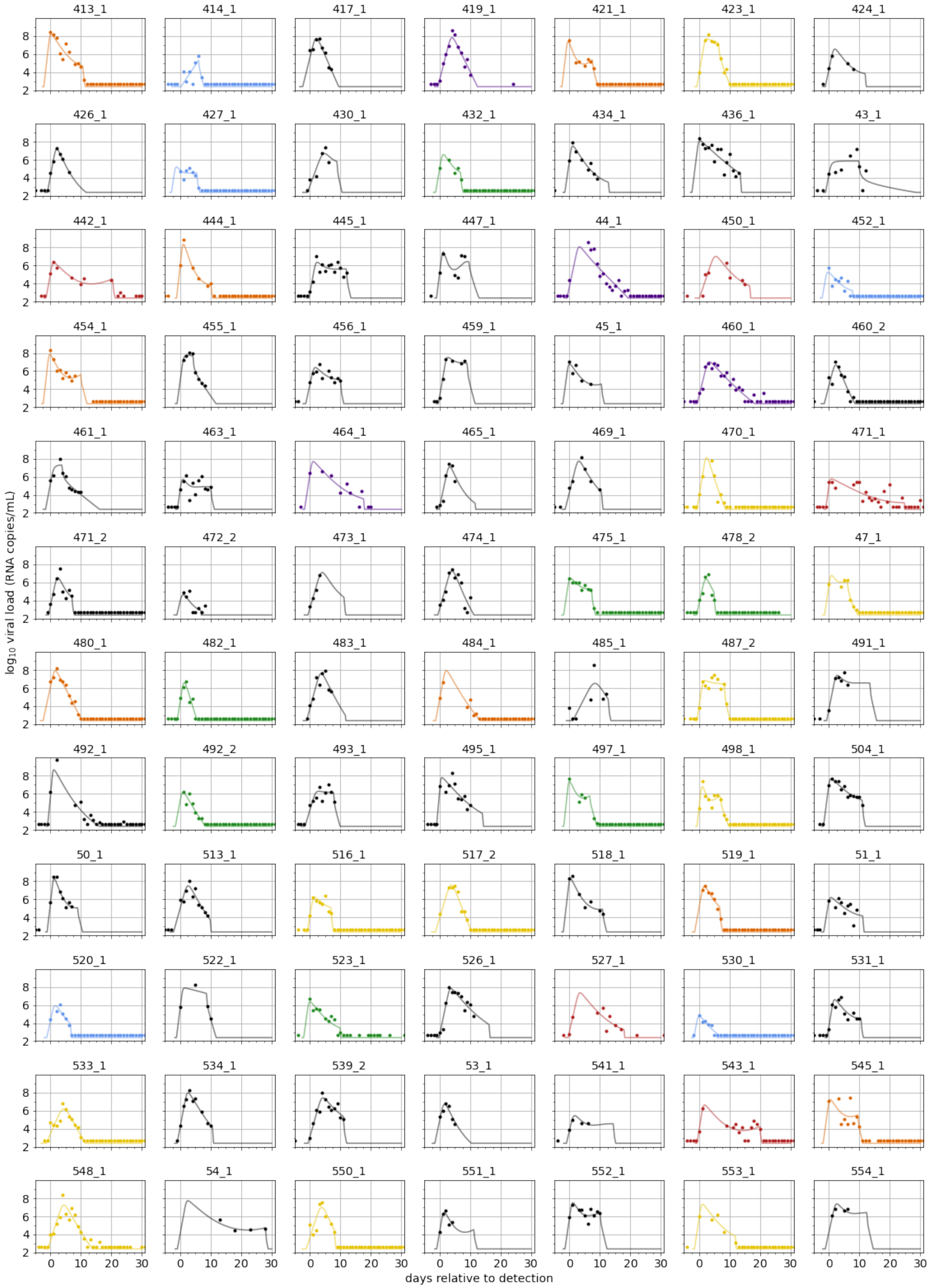
413_1
414_1
417_1
419_1
421_1
423_1
424_1
426_1
427_1
430_1
432_1
434_1
436_1
43_1
442_1
444_1
445_1
447_1
44_1
450_1
452_1
454_1
455_1
456_1
459_1
45_1
460_1
460_2
461_1
463_1
464_1
465_1
469_1
470_1
471_1
471_2
472_2
473_1
474_1
475_1
478_2
47_1
480_1
482_1
483_1
484_1
485_1
487_2
491_1
492_1
492_2
493_1
495_1
497_1
498_1
504_1
50_1
513_1
516_1
517_2
518_1
519_1
51_1
520_1
522_1
523_1
526_1
527_1
530_1
531_1
533_1
534_1
539_2
53_1
541_1
543_1
545_1
548_1
54_1
550_1
551_1
552_1
553_1
554_1
$\log_{10}$ viral load (RNA copies/mL)
days relative to detection

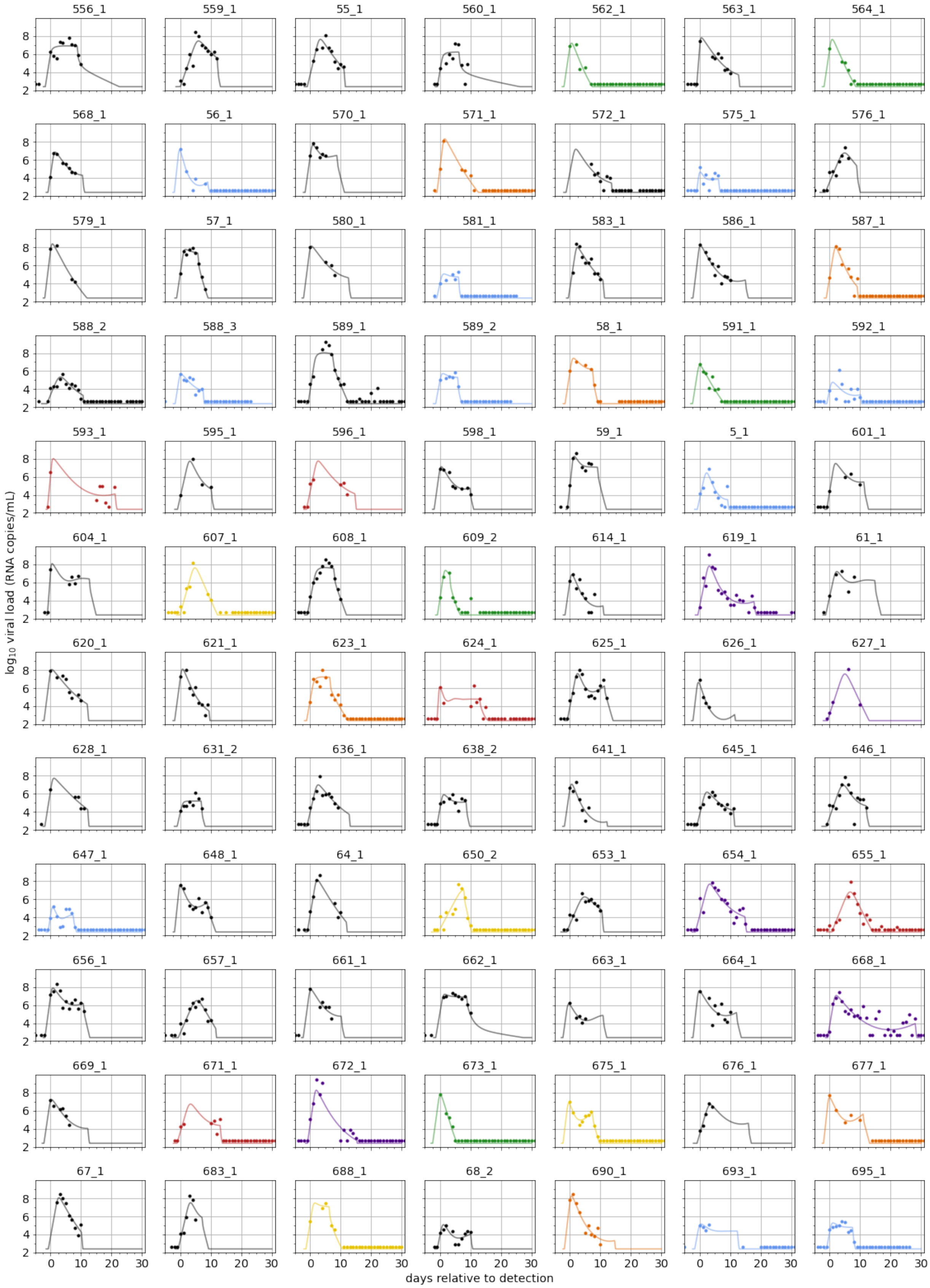
556_1
559_1
55_1
560_1
562_1
563_1
564_1
568_1
56_1
570_1
571_1
572_1
575_1
576_1
579_1
57_1
580_1
581_1
583_1
586_1
587_1
588_2
588_3
589_1
589_2
58_1
591_1
592_1
593_1
595_1
596_1
598_1
59_1
5_1
601_1
604_1
607_1
608_1
609_2
614_1
619_1
61_1
620_1
621_1
623_1
624_1
625_1
626_1
627_1
628_1
631_2
636_1
638_2
641_1
645_1
646_1
647_1
648_1
64_1
650_2
653_1
654_1
655_1
656_1
657_1
661_1
662_1
663_1
664_1
668_1
669_1
671_1
672_1
673_1
675_1
676_1
677_1
67_1
683_1
688_1
68_2
690_1
693_1
695_1
$\log_{10}$ viral load (RNA copies/mL)
days relative to detection

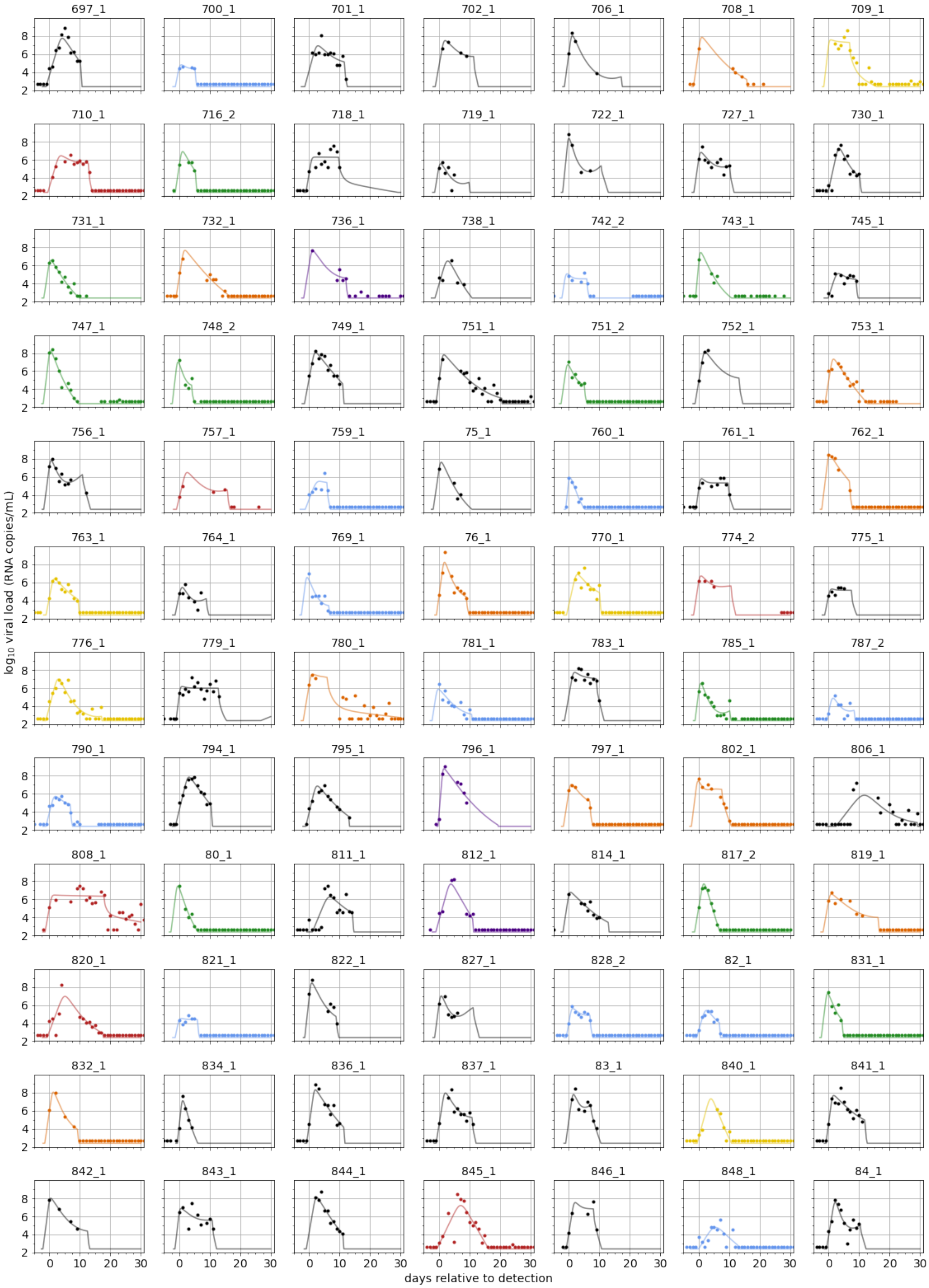
697_1
700_1
701_1
702_1
706_1
708_1
709_1
710_1
716_2
718_1
719_1
722_1
727_1
730_1
731_1
732_1
736_1
738_1
742_2
743_1
745_1
747_1
748_2
749_1
751_1
751_2
752_1
753_1
756_1
757_1
759_1
75_1
760_1
761_1
762_1
763_1
764_1
769_1
76_1
770_1
774_2
775_1
776_1
779_1
780_1
781_1
783_1
785_1
787_2
790_1
794_1
795_1
796_1
797_1
802_1
806_1
808_1
80_1
811_1
812_1
814_1
817_2
819_1
820_1
821_1
822_1
827_1
828_2
82_1
831_1
832_1
834_1
836_1
837_1
83_1
840_1
841_1
842_1
843_1
844_1
845_1
846_1
848_1
84_1
$\log_{10}$ viral load (RNA copies/mL)
days relative to detection

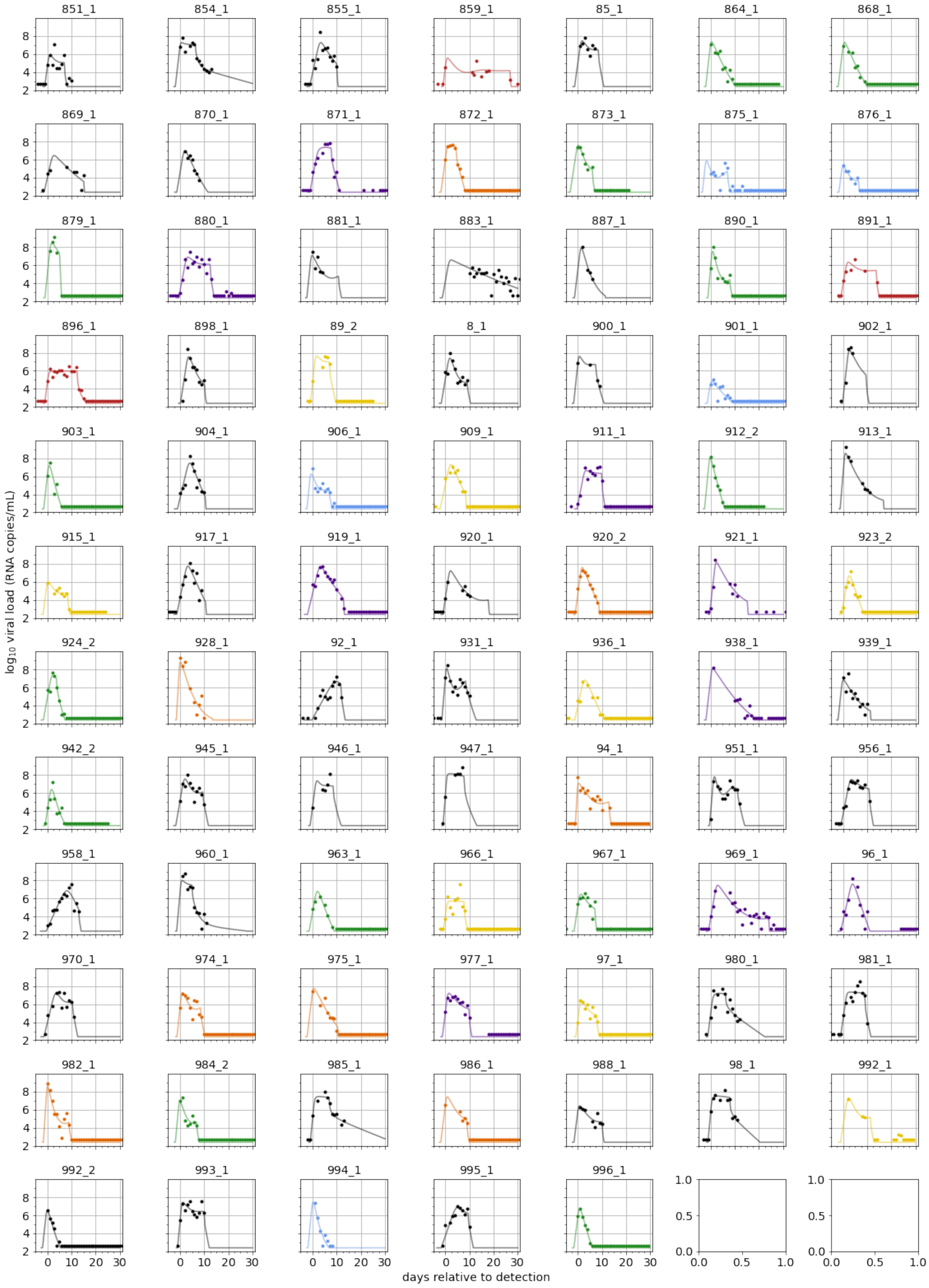

851_1
854_1
855_1
859_1
85_1
864_1
868_1
869_1
870_1
871_1
872_1
873_1
875_1
876_1
879_1
880_1
881_1
883_1
887_1
890_1
891_1
896_1
898_1
89_2
8_1
900_1
901_1
902_1
903_1
904_1
906_1
909_1
911_1
912_2
913_1
915_1
917_1
919_1
920_1
920_2
921_1
923_2
924_2
928_1
92_1
931_1
936_1
938_1
939_1
942_2
945_1
946_1
947_1
94_1
951_1
956_1
958_1
960_1
963_1
966_1
967_1
969_1
96_1
970_1
974_1
975_1
977_1
97_1
980_1
981_1
982_1
984_2
985_1
986_1
988_1
98_1
992_1
992_2
993_1
994_1
995_1
996_1
$\log_{10}$ viral load (RNA copies/mL)
days relative to detection
